# A fully remote randomized controlled trial of an ultra-brief digital meditation intervention reduces internalizing symptoms

**DOI:** 10.64898/2026.04.19.26351219

**Authors:** Cameron C. Glick, Saad Pirzada, Shaun Quah, Sasha Feldman, Iyọbosa Enabulele, Soren Madsen, Neel Billimoria, Summer Feldman, Richa Bhatia, David Spiegel, Manish Saggar

## Abstract

**Background:** Scalable, low-burden behavioral interventions are needed to address rising subclinical mental health symptoms. However, few randomized controlled trials have evaluated ultra-brief, remotely delivered, meditation using multimodal outcome assessment under real-world conditions.

**Methods:** We conducted a fully remote randomized controlled trial (ClinicalTrials.gov: NCT06014281) evaluating a focused-attention meditation intervention delivered via brief instructor training and independent daily practice. A total of 299 meditation-naïve adults were randomized to immediate intervention or waitlist control in a delayed-intervention design. Participants practiced ≥10 minutes daily for 8 weeks within a 16-week study. Outcomes included validated self-report measures, web-based cognitive tasks, and wearable-derived physiological metrics.

**Results:** Across randomized and within-participant replication phases, the intervention was associated with significant reductions in anxiety and mind wandering, with effects remaining stable during 8-week follow-up. Improvements were greatest among participants with higher baseline symptom burden. Sleep disturbance improved selectively among individuals with poorer baseline sleep. Secondary outcomes, including rumination, perceived stress, social connectedness, and quality of life, also improved. Cognitive performance showed modest improvements primarily among lower-performing participants. Resting heart rate exhibited nominal reductions.

**Conclusions:** An ultra-brief, fully remote meditation intervention requiring 10 minutes per day was associated with sustained improvements in psychological functioning and smaller, baseline-dependent effects on cognition in a non-clinical population. These findings support digital delivery of low-dose meditation as a scalable preventive mental health strategy.

## Introduction

The COVID-19 pandemic has led to a sustained global increase in stress, anxiety, and depressive symptoms across diverse populations, including individuals with no prior psychiatric history (Bourmistrova et al., 2022; Hossain et al., 2020; Kumar & Nayar, 2021; Liu et al., 2020; Saltzman et al., 2024; Shanbehzadeh et al., 2021). Large-scale epidemiological studies have documented persistent elevations in psychological distress well beyond the acute phases of lockdowns, with many individuals reporting subclinical yet functionally impairing levels of stress and anxiety (COVID-19 Mental Disorders Collaborators, 2021; Pierce et al., 2020; Touron et al., 2025). Although these symptoms do not meet diagnostic criteria for psychiatric disorders, they are associated with reduced well-being, impaired sleep, decreased productivity, and increased risk of future psychopathology (Cuijpers & Smit, 2004; Konstantopoulou et al., 2020). Because individuals with subthreshold symptoms often fall outside traditional treatment pathways, scalable preventive interventions are urgently needed.

Meditation and related attentional training practices represent promising low-cost approaches for improving emotional regulation and reducing stress. Meta-analyses demonstrate moderate reductions in anxiety, stress, and depressive symptoms across clinical and non-clinical populations (Goldberg et al., 2022; Goyal et al., 2014; Khoury et al., 2013), potentially mediated by improvements in attentional control and regulation of stress-related physiological processes (Fox et al., 2016; Saggar et al., 2012, 2015; Tang et al., 2015). However, most evidence derives from intensive in-person training programs or time-demanding daily practices, limiting real-world accessibility and long-term adherence.

Digital mental-health interventions provide an opportunity to overcome these barriers. Mobile-health approaches allow individuals to engage in behavioral interventions remotely, at their own pace, and with minimal infrastructure (Firth et al., 2017; Torous et al., 2021). For stress and anxiety management in particular, sustained daily practice may be more important than session intensity, suggesting that brief, repeatable interventions could offer scalable benefits (Elbaz et al., 2021; Staiano et al., 2025). Yet few randomized controlled trials have tested whether very short daily meditation practices, on the order of minutes rather than tens of minutes, can produce measurable psychological or physiological effects.

The present study evaluates a focused-attention meditation practice commonly referred to as SOS meditation. Historically SOS meditation is taught within a broader contemplative tradition but implemented here as a secular attentional training exercise (Munjal et al., 2025; Singh, 2022). The practice involves directing attention inward while maintaining relaxed awareness and repeating a neutral word or phrase. Unlike many structured mindfulness programs, the technique requires minimal instruction, no specialized posture, and no ongoing instructor involvement, making it suitable for remote delivery and routine integration into daily life. From a theoretical perspective, the practice aligns with neurocognitive models of mindfulness emphasizing inward attention, narrow attentional aperture, and meta-awareness (Lutz et al., 2015). Repeated engagement with these attentional processes may, over time, reduce rumination and improve awareness of ongoing cognitive states while imposing relatively low cognitive effort, potentially supporting adherence among meditation-naïve individuals. Preliminary reports suggest beneficial psychological effects in remote settings (Munjal et al., 2025), but its broader cognitive and physiological impacts, particularly at brief daily durations, remain unclear.

To address methodological limitations of prior meditation research, including small samples, uncontrolled designs, and reliance only on self-report outcomes (Goldberg et al., 2022; Van Dam et al., 2018), we conducted a fully remote randomized controlled trial with delayed-intervention replication. Participants were assigned to either an immediate intervention group or a waitlist control group that later received the same training, enabling both between-group comparisons and within-participant replication. We recruited 299 meditation-naïve adults to complete a 16-week study consisting of an 8-week intervention and follow-up period. Participants practiced at least 10 minutes per day using a remotely delivered training protocol.

To capture potential effects across multiple systems, we adopted a multimodal assessment framework. Psychological outcomes included validated measures of generalized anxiety (GAD-7), sleep disturbance (PSQI), and mind wandering (MWQ), along with secondary measures of perceived stress, depressive symptoms, rumination, social connectedness, quality of life, and character strengths. Cognitive performance was assessed using web-based executive function and working memory tasks (Stroop and 2-back). Physiological indices included resting heart rate and heart rate variability derived from nightly wearable recordings. This integrative design allowed us to examine whether brief daily meditation practice produces convergent changes across subjective experience, cognitive performance, and autonomic regulation.

We hypothesized that a brief, remotely delivered focused-attention meditation intervention would reduce anxiety and stress-related symptoms and produce convergent improvements across psychological, cognitive, and physiological domains. By testing a minimal-burden intervention under real-world conditions, this study evaluates the feasibility and efficacy of ultra-brief daily meditation as a scalable preventive mental-health strategy.

## Results

### Study Population

A total of 681 individuals were assessed for eligibility, of whom 299 were randomized to either the immediate intervention group (Arm 1, n=151) or the waitlist control group (Arm 2, n=148). Participant flow, attrition, and analysis inclusion are summarized in the CONSORT diagram (Fig. 1).

**Fig. 1.**
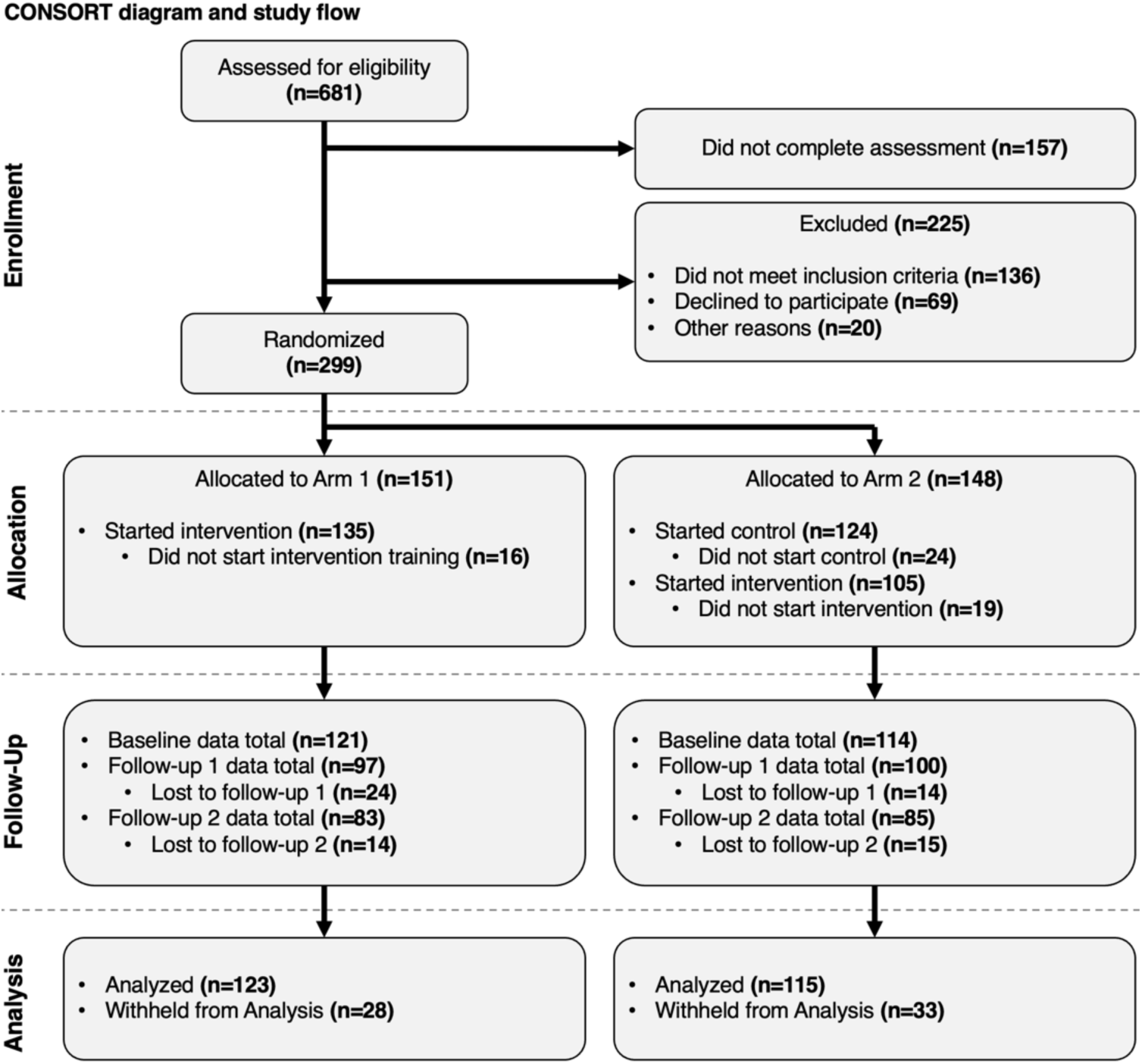
CONSORT flow diagram of participant recruitment, allocation, follow-up, and analysis. A total of 681 individuals were assessed for eligibility, of whom 299 were randomized to either the immediate intervention arm (n=151) or the waitlist control arm (n=148). The diagram summarizes exclusions, intervention initiation, follow-up completion, and inclusion in analysis for each arm.

The primary analysis sample had a mean age of 33.8 years and was 66.7% female. Baseline demographic and clinical characteristics were similar across study arms (Table 1). Primary analyses adjusted for age and sex.

**Table 1.**
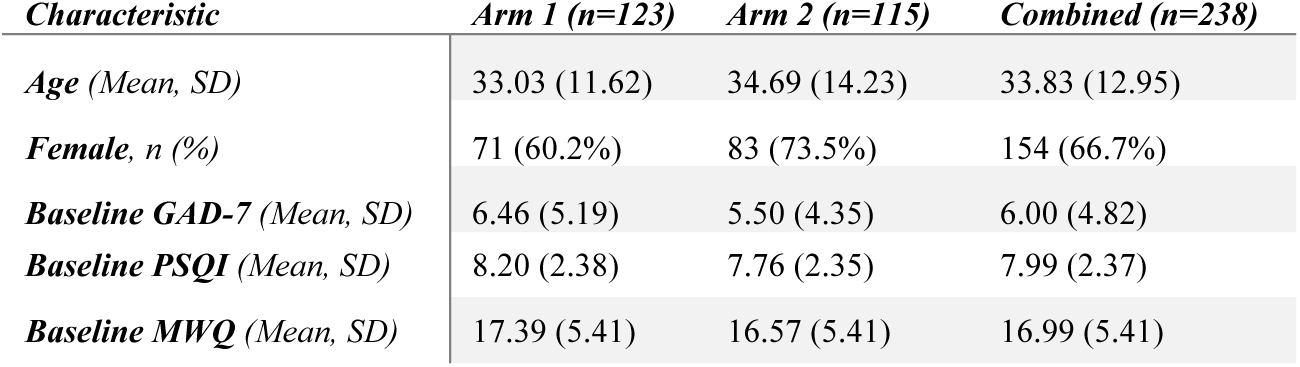
Baseline characteristics of participants included in the primary analysis set. Values are mean (SD) or n (%). All analyses adjusted for age and sex.

| Characteristic | Arm 1 (n=123) | Arm 2 (n=115) | Combined (n=238) |
| --- | --- | --- | --- |
| Age (Mean, SD) | 33.03 (11.62) | 34.69 (14.23) | 33.83 (12.95) |
| Female, n (%) | 71 (60.2%) | 83 (73.5%) | 154 (66.7%) |
| Baseline GAD-7 (Mean, SD) | 6.46 (5.19) | 5.50 (4.35) | 6.00 (4.82) |
| Baseline PSQI (Mean, SD) | 8.20 (2.38) | 7.76 (2.35) | 7.99 (2.37) |
| Baseline MWQ (Mean, SD) | 17.39 (5.41) | 16.57 (5.41) | 16.99 (5.41) |

### Anxiety and mind wandering decrease following brief daily meditation, particularly among those with elevated baseline symptoms

#### Anxiety decreased following meditation intervention and effects were sustained

Daily meditation was associated with significant reductions in anxiety symptoms, as measured by the GAD-7 (Fig. 2ai). During the randomized phase (T1–T2), participants assigned to the immediate intervention arm exhibited greater reductions in anxiety relative to the waitlist control arm (F=4.989, *p*=0.038; Table S1). When the waitlist group subsequently received the intervention (T2–T3), a comparable within-participant reduction was observed (t=-3.849, *p*<0.001; Table S2), providing internal replication of the effect. Anxiety reductions were maintained at follow-up, with no evidence of rebound (Table S3). FDR correction was used in all analyses to control for multiple comparisons.

**Fig. 2.**
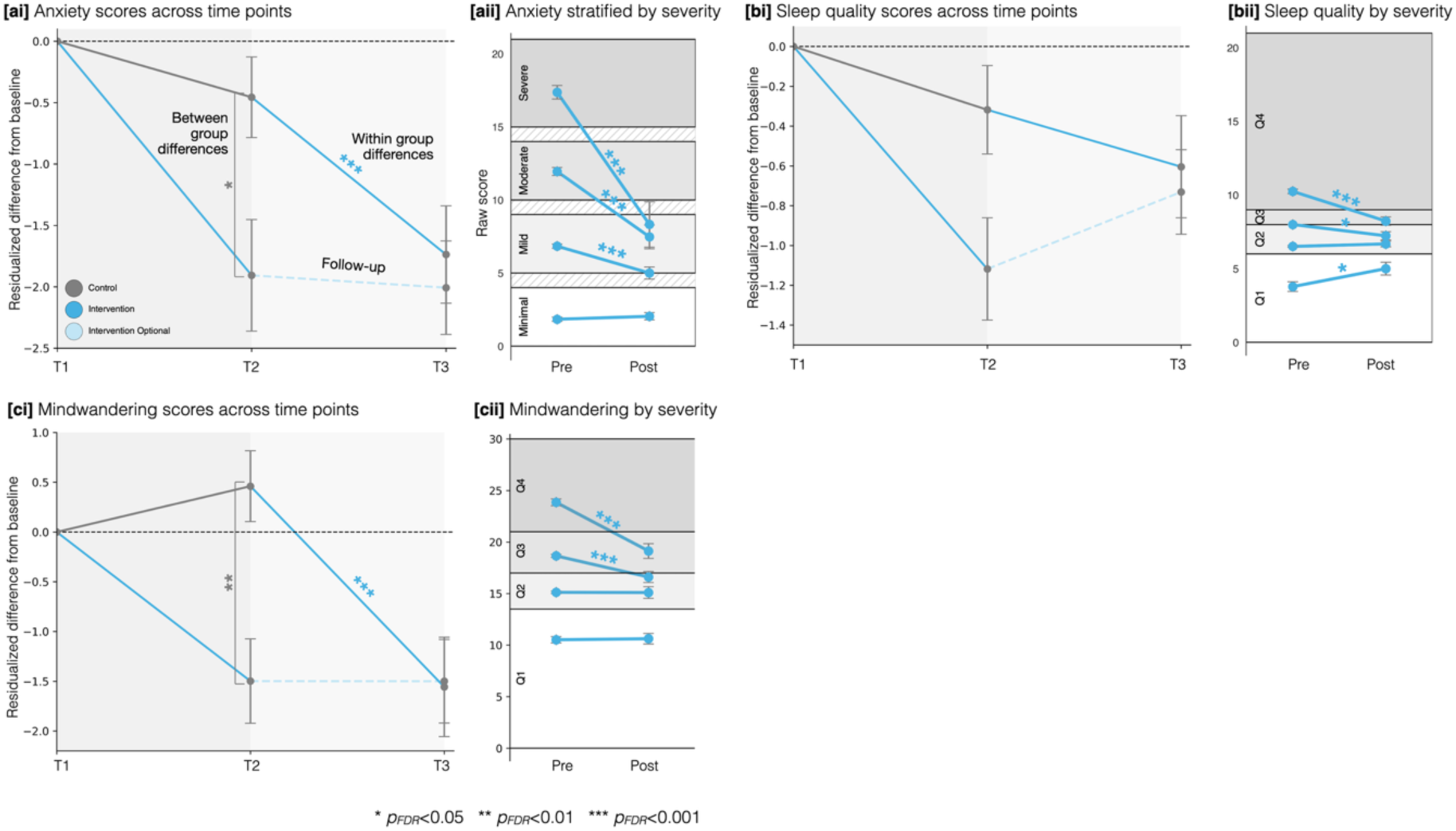
Primary psychological outcomes. [ai-ci] Residualized change from baseline in anxiety (GAD-7), sleep disturbance (PSQI), and mind wandering (MWQ) across study timepoints. During the randomized phase (T1–T2), participants assigned to the intervention arm showed greater reductions in anxiety and mind wandering relative to the waitlist control arm. Effects were replicated when the waitlist group subsequently received the intervention (T2–T3) and were sustained at follow-up. Error bars represent SEM. [aii-cii] Pre–post changes stratified by baseline severity are also presented. Participants with higher baseline symptom levels exhibited larger reductions following the intervention, whereas individuals with minimal baseline symptoms showed little change. Significance markers reflect FDR-adjusted tests.

Pooling intervention periods across both arms revealed that participants with moderate or severe baseline anxiety showed the largest improvements. Individuals with baseline GAD-7 scores (10–14) or (15-21) demonstrated marked reductions (both *p*<0.0001; Table S4), with many shifting to lower clinical severity categories following the intervention (Fig. 2aii). Participants with minimal baseline anxiety showed little change.

#### Sleep improved selectively among participants with baseline disturbance

Across the three study points, changes in sleep disturbance (measured using the PSQI) did not reach statistical significance (Fig. 2bi; Tables S1–S3). However, stratified analyses revealed significant improvements among participants with third and fourth quartile baseline sleep disturbance (*p*<0.05; Fig. 2bii; Table S4), whereas individuals with first quartile baseline sleep showed somewhat higher PSQI scores post-intervention (p=0.042; Table S4).

These findings suggest that sleep-related effects of brief meditation may be contingent upon baseline symptom burden.

#### Mind wandering decreased and effects were sustained

Similar to anxiety scores, mind wandering (assessed using the MWQ) also declined following daily meditation (Fig. 2ci). During the randomized phase, intervention participants showed greater reductions relative to controls (F=9.817, *p*=0.005; Table S1). Replication was observed when the waitlist group received training (t=-4.146, *p*<0.001; Table S2). Reductions were sustained eight weeks after the intervention period.

Similar to anxiety, participants in the highest baseline quartiles of mind wandering exhibited the largest decreases (Q3 and Q4, both *p*<0.001; Fig. 2cii; Table S4), whereas individuals with low baseline levels showed no change.

Across the three primary self-report psychological outcomes, meditation was associated with robust reductions in anxiety and mind wandering, with sustained effects at follow-up. Sleep improvements were baseline dependent. In all domains, individuals with higher baseline symptom severity exhibited the largest gains.

### Cognitive improvements emerge primarily among lower-performing participants

Cognitive performance was assessed using repeated administrations of the Stroop and 2-back working memory tasks. At the whole-sample level, no significant between-arm differences in accuracy were observed during the randomized phase (Fig. 3ai-di; Tables S9–S11). Response times decreased over time across both intervention and control arms, consistent with task familiarity or practice effects, and did not yield robust intervention-specific effects.

**Fig. 3.**
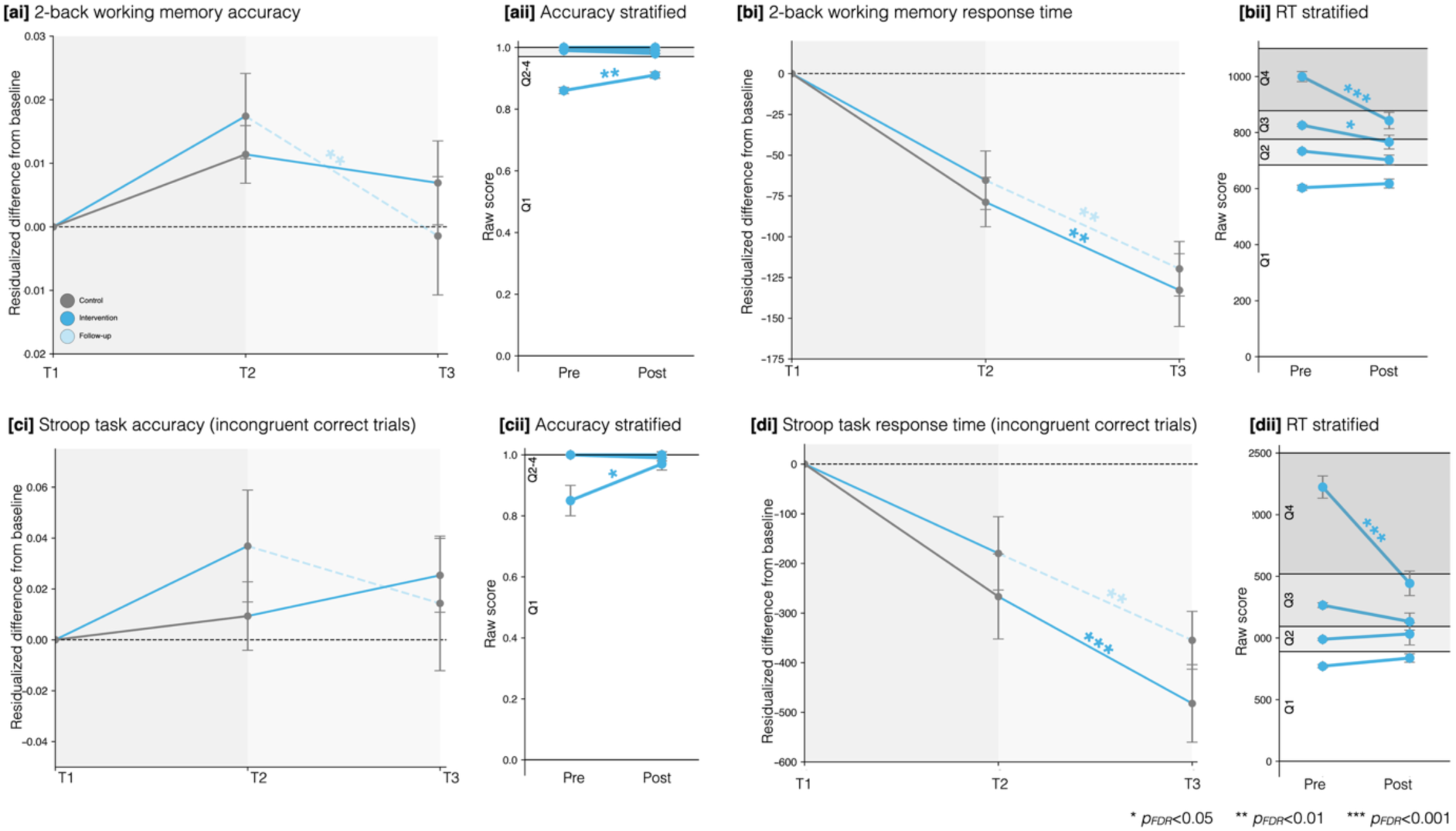
Primary cognitive outcomes. (ai-di) Residualized change from baseline in 2-back accuracy, 2-back reaction time, Stroop incongruent accuracy, and Stroop incongruent response time across study timepoints. No robust between-arm differences were observed at the whole-sample level, with improvements in reaction time occurring across both intervention and control periods. Pre–post changes stratified by baseline performance quartile are also presented. (aii-dii) Participants in the lowest-performing quartile at baseline exhibited significant improvements in accuracy and reaction time following meditation exposure, whereas higher-performing participants showed minimal change. Error bars represent SEM. Significance markers reflect FDR-adjusted tests.

Baseline inspection revealed pronounced ceiling effects in accuracy across both tasks. To evaluate potential benefits among individuals with greater room for improvement, we conducted stratified analyses. Within the lowest-performing quartile, at baseline, participants exposed to meditation demonstrated significant improvement in accuracy and reaction time on both the Stroop and 2-back tasks (both *p*<0.05; Fig. 3aii-dii; Table S12). This finding suggests that brief daily meditation may confer modest cognitive benefits in participants with lower baseline executive performance, although robust group-level effects were not observed.

### Autonomic measures show directional but non-significant changes

Physiological indices were assessed using wearable-derived resting heart rate (RHR) and heart rate variability (HRV) collected throughout the study period. During the randomized phase, RHR showed a consistent downward trend among participants engaged in meditation relative to the waitlist control arm; however, between-group differences did not reach statistical significance (Fig. S1; Tables S13–S16). When the waitlist group subsequently received the intervention, an insignificant directional reduction in RHR was observed. Among initial intervention participants, reductions were maintained through follow-up.

Similarly, no significant group-level changes were detected for HRV across randomized or within-group phases. HRV exhibited substantial inter-individual variability and did not differentiate intervention from control periods.

### Broad improvements across connectedness, rumination, stress, and quality of life

Secondary psychological measures showed patterns broadly consistent with primary outcomes (Fig. 4). Although between-arm differences at week 8 did not survive FDR correction, nominal trends (uncorrected *p*<0.05) were observed for social connectedness, rumination, and quality of life scores (Table S5). Further, within-participant quartile-based analyses following meditation exposure also revealed significant improvements across several domains (Tables S6–S8).

**Fig. 4.**
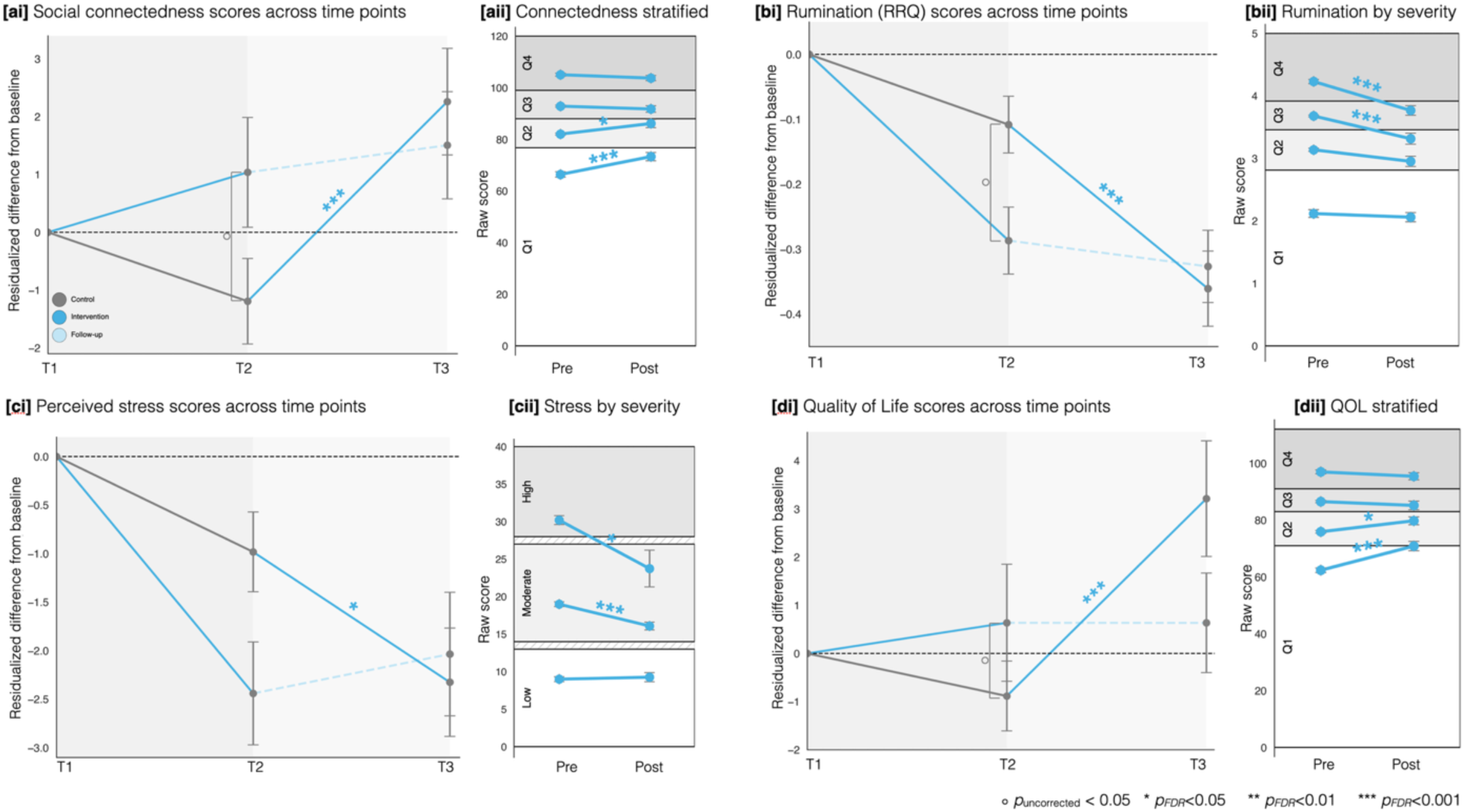
Secondary psychological outcomes. (a-d) Residualized change from baseline in secondary self-report measures, including social connectedness, rumination, perceived stress, and quality of life, across study timepoints. Group-level effects were directionally consistent with primary outcomes but generally smaller in magnitude and nominally significant group differences (uncorrected p < 0.05). Pre–post changes stratified by baseline severity or quartiles are also presented. Participants with lower baseline connectedness, higher rumination or stress, and lower quality of life exhibited the largest improvements following meditation exposure, whereas participants with minimal baseline impairment showed little change. Error bars represent SEM. Significance markers indicate FDR-adjusted tests.

#### Social connectedness

Social connectedness increased during meditation periods and replicated in the delayed-intervention arm (t=3.729, *p*<0.001; Table S6). Participants with lower baseline connectedness exhibited the largest gains (Fig. 4ai-ii; Table S8).

#### Rumination

Rumination decreased following meditation exposure in both arms. Reductions were most pronounced among individuals with elevated baseline rumination (Fig. 4bi-ii; upper quartiles; *p*<0.001; Table S8), consistent with a shift in maladaptive repetitive thought patterns.

#### Perceived stress

Perceived stress showed directional reductions across arms, reaching significance during the delayed-intervention phase (Fig. 4ci-ii; t=-2.496, *p*=0.023; Table S6), suggesting modest stress-related benefits.

#### Quality of life

Quality of life improved following meditation, particularly among participants reporting lower baseline well-being (Fig. 4di-ii; worst quartile; t=5.335, p<0.001; Table S8).

The remaining secondary psychological measures showed a more selective and baseline-dependent pattern. Depressive symptoms improved mainly among participants with elevated baseline PHQ-8 scores, and the worst quartile for reflection showed an increase following meditation exposure, whereas the BSS-12 subscales exhibited mixed changes that were not consistently supported at the quartile level after correction for multiple comparisons.

Complete statistical details are provided in Supplementary Tables S5–S8.

### Summary of univariate analysis

Across the psychological, cognitive, and physiological domains, the most robust and sustained effects were observed in primary psychological outcomes, with baseline-dependent effects in cognition and modest, non-significant trends in physiological measures. A summary of the primary imputed univariate results is provided in Table 2; corresponding non-imputed results are reported in Table S17. The overall pattern of findings was consistent with or without imputation.

**Table 2.**
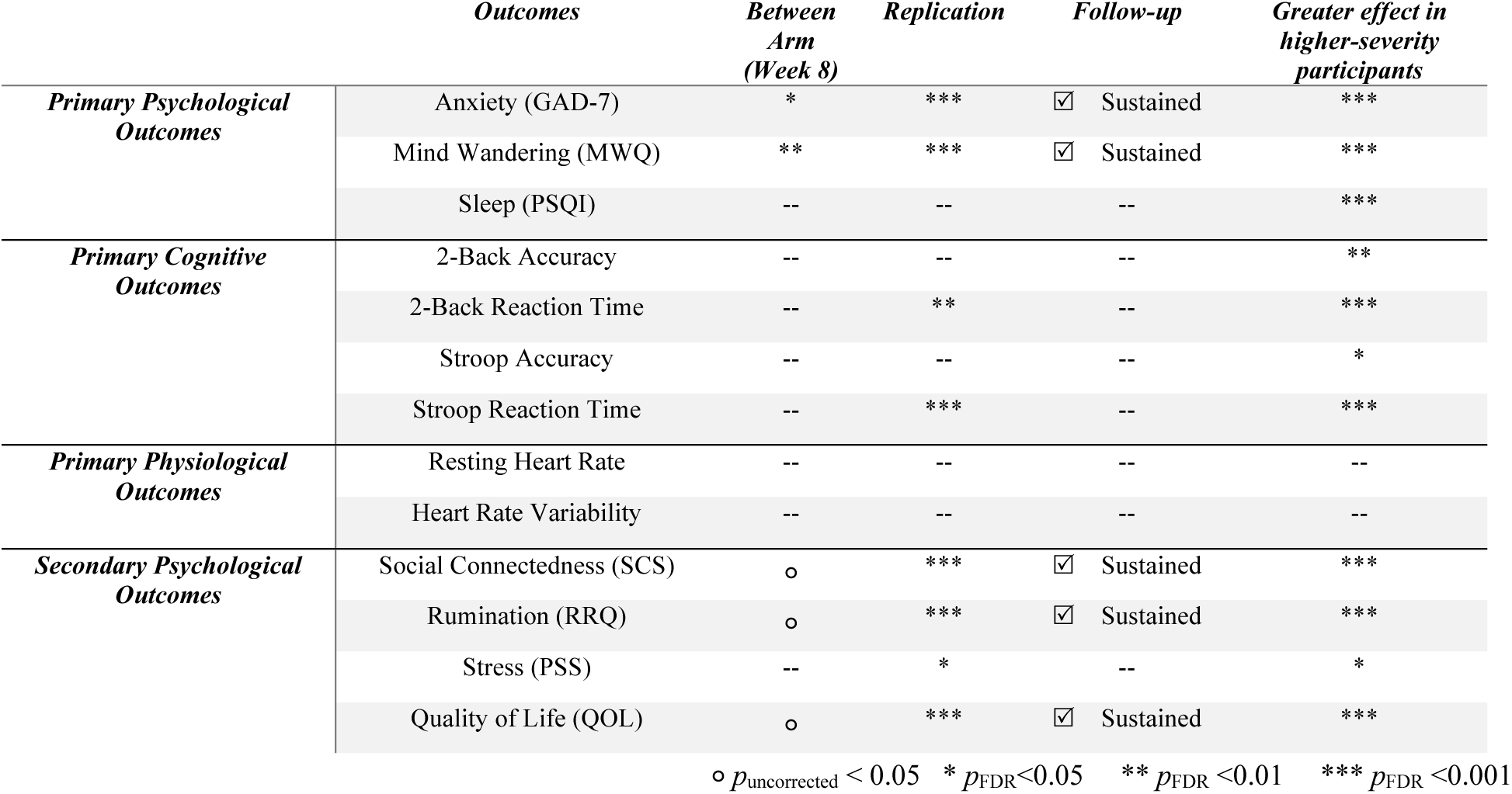
Consolidated summary of univariate analysis. Intervention effects across all the primary and secondary outcomes. Between-arm (T2; Week 8) reflects the randomized comparison between intervention and waitlist control groups. Replication reflects within-arm change in the delayed-intervention group following meditation exposure. Follow-up reflects stability of change from Week 8 (T2) to Week 16 (T3) in the initial intervention arm. Greater effect in higher-severity participants reflects baseline-stratified analyses among participants in the highest symptom quartile or clinically elevated range. Significance markers denote FDR-corrected p-values.

### Multivariate Structure of Self-Reported Measures

#### Self-reported measures are organized along two latent dimensions

To characterize shared variance across self-reported psychological measures, we performed exploratory factor analysis (EFA) on baseline questionnaire scores. A two-factor solution emerged, accounting for a substantial proportion of total variance (42%) (Fig 5a; Fig. S2; Table S18). Factor loadings were thresholded at 0.40 to enhance interpretability, consistent with established recommendations for interpretability (Brooks & Stevens, 1994), and factors were named based on the highest-loading measures.

**Fig. 5.**
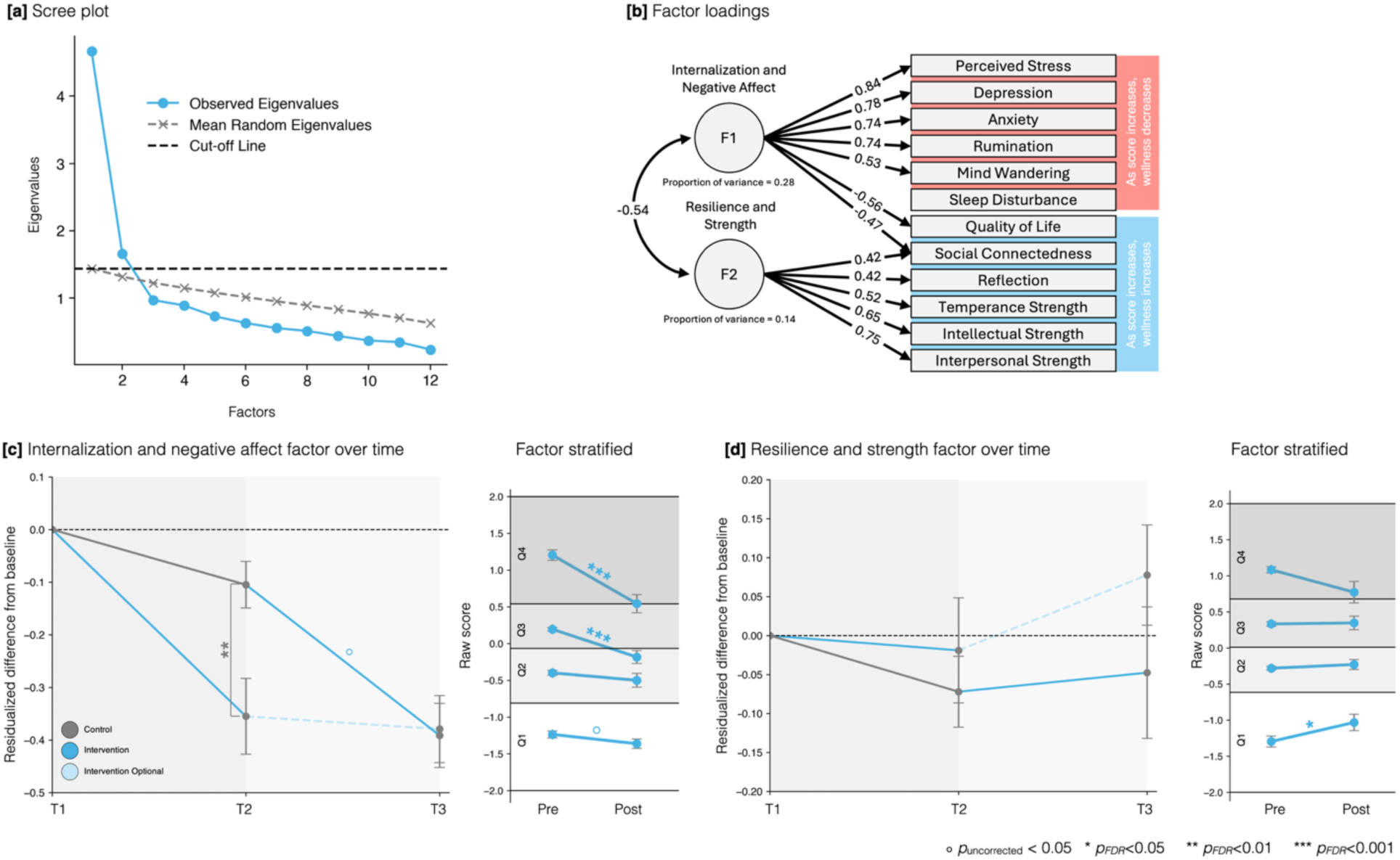
Exploratory factor analysis and latent change following meditation. (a) Scree plot and parallel analysis of baseline (T1) self-report measures indicating a two-factor solution. The dashed line represents mean eigenvalues derived from 1,000 random permutations. (b) Factor loadings (|loading| ≥ 0.40) for the two-factor model fitted on baseline data. Factor 1 (Internalization and Negative Affect) and Factor 2 (Resilience and Strength) were moderately negatively correlated (r = -0.54). (c) Change in Internalization and Negative Affect factor scores across study timepoints. Residualized change from baseline is shown for Arm 1 (intervention) and Arm 2 (control), with within-participant replication (Arm 2, T2–T3) and follow-up (Arm 1, T2–T3) phases. Right panel shows pre–post changes stratified by baseline factor quartiles, demonstrating greater reductions among participants with higher baseline scores. (d) Change in Resilience and Strength factor scores across study timepoints, displayed analogously to (c).

The first factor, termed *Internalization and Negative Affect*, was defined by strong positive loadings from perceived stress, depressive symptoms, anxiety, rumination, and mind wandering, and negative loadings from quality of life and social connectedness. The second factor, termed *Resilience and Strength*, was characterized by positive loadings from intellectual, interpersonal, and temperance character strengths, as well as reflection. The two factors were moderately negatively correlated, indicating related but distinct psychological dimensions (r = -0.54, *p*<0.05; Fig. 5b).

#### Meditation selectively reduces the latent internalization dimension

Factor scores derived from the baseline-fitted EFA model were examined across study timepoints using the same analytic framework as univariate outcomes. For the Internalization and Negative Affect factor, we observed a significant reduction during the randomized phase (T1–T2) in the intervention arm relative to control (F=11.195, *p*=0.002; Table S19), with consistent within-participant replication in Arm 2 during the delayed intervention period (T2–T3; t=-5.106, *p*<0.001; Table S20) and sustained effects in Arm 1 at follow-up. These results indicate a robust shift along the latent symptom axis of internalization following meditation (Fig. 5c). Baseline-stratified analyses revealed that participants with the highest symptom burden benefited the most from meditation intervention (Table S22).

In contrast, changes in the Resilience and Strength factor were not significant at the whole-sample level and did not show consistent between-arm differences during the randomized phase. However, baseline-stratified analyses revealed differential effects, with participants in the lowest baseline quartile exhibiting significant increases in resilience-related scores following intervention, whereas higher-baseline groups showed no change (t=2.281, *p*=0.02; Table S22).

Together, these findings suggest that the multivariate effects of brief daily meditation are primarily expressed as reductions in internalizing symptomatology, with more selective and baseline-dependent changes in resilience-related traits.

### Baseline latent factors associated with individual response across domains

Although meditation reduced Internalization and Negative Affect at the group level, substantial inter-individual variability in change scores was observed. To examine predictors of treatment response, we computed partial correlations between baseline factor scores and post–pre change in individual outcomes, controlling for age and sex. Analyses were conducted separately across psychological, physiological, and cognitive domains with FDR correction applied within each domain.

#### Associations with psychological outcomes

Higher baseline Internalization and Negative Affect scores were associated with larger reductions in anxiety (r=-0.245, *p*=0.0123) and larger increases in quality of life (r=0.291, *p*=0.002) and social connectedness (r=0.310, *p*=0.002; Table S23). Associations with perceived stress and depressive symptoms maintained a similar direction and were nominal at the raw level but did not survive FDR correction.

These findings indicate that individuals with elevated internalizing symptom burden at baseline experienced greater psychological benefit from brief daily meditation practice.

Baseline Resilience and Strength scores showed a different pattern. Higher resilience was associated with reductions in social connectedness following the intervention (r = -0.250, *p* = 0.0121).

#### Associations with cognitive and physiological outcomes

No significant associations were observed between baseline latent factor scores and changes in changes in cognitive performance or physiological measures (Table S22-23).

## Discussion

This randomized, fully remote trial demonstrates that a brief (10 minutes/day) digitally delivered focused-attention meditation intervention can produce measurable improvements in psychological functioning, with effects concentrated along a latent internalizing symptom dimension and strongest among individuals with elevated baseline symptom burden. By integrating validated self-report psychological measures, web-based cognitive testing, and wearable-derived physiological indices within a scalable remote design, this study advances prior internet-based mindfulness trials that have typically relied on single-domain outcomes or lacked multimodal assessment (Devillers-Réolon et al., 2022; Sevilla-Llewellyn-Jones et al., 2018; Simonsson & Marks, 2020).

### Psychological Outcomes

Self-report measures demonstrated robust and consistent psychological benefits of brief regular meditation practice. Participants in both study arms exhibited reductions in anxiety and mind wandering, with the largest improvements observed among individuals who entered the study with moderate to severe baseline symptom levels. Sleep disturbance showed a more nuanced pattern, with improvements primarily evident among participants with the worst quartile of sleep difficulties, while individuals with the best quartile of baseline sleep showed higher PSQI scores post-intervention. These findings are consistent with prior work demonstrating that meditation interventions tend to be most effective for individuals with elevated symptom burden (Kabat-Zinn et al., 1992; Krisanaprakornkit et al., 2006; Orme-Johnson & Barnes, 2014).

Importantly, reductions in anxiety and mind wandering remained stable during the follow-up period, with no evidence of symptom rebound. These sustained/stable effects suggest that brief SOS meditation may induce changes in attentional and affective processes that generalize beyond periods of active practice.

The observed reductions in mind wandering align with prior neurocognitive and phenomenological work linking meditation to enhanced meta-awareness and attentional control (Hasenkamp et al., 2012; Rahl et al., 2017). For anxiety, the magnitude of change on the GAD-7 fell within ranges identified as minimally clinically important, underscoring the clinical relevance of the findings beyond statistical significance.

These univariate improvements were mirrored at the latent level, where meditation was associated with a reduction along a shared Internalization and Negative Affect dimension.

### Cognitive Outcomes

Cognitive effects of a brief daily meditation were more modest. At the whole-sample level, no robust between-group differences were observed on Stroop or 2-back task performance, likely reflecting ceiling effects in this high-functioning, non-clinical sample. However, among participants in the worst-performing quartile at baseline, meditation was associated with improvements in accuracy and processing speed, particularly on Stroop incongruent trials. These findings are consistent with recent meta-analytic work suggesting small but detectable benefits of mindfulness-related practices on working memory and cognitive control, especially among individuals with lower baseline performance (Zainal & Newman, 2024).

The modest effect sizes observed here are also consistent with the broader literature, which remains mixed regarding meditation’s impact on cognition (Demnitz-King et al., 2023; Zainal & Newman, 2024). Several factors may have constrained detectable cognitive effects in the present study, including the brief duration of the intervention, individual variability in cognitive strategies, and limitations inherent to remote testing environments (e.g., device heterogeneity, learning effects from repeated task exposure). Nonetheless, the subgroup-specific improvements observed here suggest that baseline performance may constrain detectable group-level effects and influence responsiveness to brief attentional training.

### Physiological Outcomes

Physiological measures derived from wearable devices revealed directionally consistent changes. Resting heart rate showed small reductions during meditation periods, with trends persisting into follow-up, though these effects did not reach statistical significance at the whole-sample level. While non-transcendental meditation practices have been associated with cardiovascular benefits in prior work (Shi et al., 2017), the predominantly healthy nature of the present sample and the brevity of the intervention likely limited the magnitude of detectable autonomic effects. Studies in clinical or higher-risk populations may reveal stronger physiological responses. Lastly, wearable-derived measures, while scalable, may lack sensitivity to subtle autonomic shifts induced by ultra-brief interventions.

### Multivariate Perspective and Individual Differences

Exploratory factor analysis revealed that participants’ self-reported experiences could be parsimoniously represented along two latent dimensions: Internalization and Negative Affect, and Resilience and Strength. Meditation was associated with a significant reduction along the Internalization and Negative Affect axis, reflecting improvements across anxiety, depression, stress, rumination, and mind wandering. Notably, baseline factor scores associated with intervention responsiveness: individuals with higher Internalization and Negative Affect showed greater psychological improvement. These findings suggest that brief digital meditation does not exert uniform effects but instead shifts individuals along a dimensional symptom continuum, with baseline position influencing magnitude and domain of response.

This pattern aligns with prior evidence suggesting that baseline characteristics meaningfully shape response to mindfulness interventions, with greater gains often observed among those with lower baseline well-being or mindfulness (Vergara et al., 2022). Together, these findings highlight the importance of personalized approaches to meditation-based interventions, where baseline psychological profiles may help identify individuals most likely to benefit across different outcome domains.

Because baseline symptom levels were correlated with magnitude of change, regression-to-the-mean effects cannot be fully excluded. However, the presence of randomized between-arm effects during the initial intervention phase and replication of improvements in the delayed intervention arm provide support for intervention-related change beyond statistical artifact.

### Limitations and Future Directions

Several limitations should be considered when interpreting the findings of this study.

#### Attrition and engagement

Attrition was substantial, with many participants failing to complete all study procedures. Commonly reported reasons included competing demands, loss of interest, and difficulty maintaining engagement over the study duration. High dropout rates are a well-documented challenge in internet-based meditation and mindfulness trials (Krisanaprakornkit et al., 2006) and underscore that self-directed meditation may not be a universally suitable intervention, particularly for individuals with lower intrinsic motivation or limited capacity for sustained self-regulation. As such, the present findings may preferentially reflect outcomes among participants who were able to remain engaged with a self-guided intervention.

#### Challenges inherent to fully remote study designs

While the fully remote nature of the trial enabled scalability and ecological validity, it also introduced several sources of data loss and variability. Participants frequently experienced difficulties with survey completion, wearable adherence, and connectivity or synchronization issues with Fitbit devices, resulting in a reduced number of complete physiological datasets. Similarly, completion rates for web-based cognitive tasks were lower than anticipated, reflecting both participant burden and the learning curve associated with unfamiliar testing platforms. In addition, remote cognitive testing is inherently sensitive to uncontrolled environmental factors (e.g., distractions, device heterogeneity, internet latency), which may have increased measurement noise and attenuated detectable effects.

#### Ceiling effects and task sensitivity

The cognitive tasks employed (Stroop and 2-back) exhibited pronounced ceiling effects in this high-functioning, non-clinical sample, limiting sensitivity to intervention-related changes at the whole-sample level. While subgroup analyses revealed improvements among participants with lower baseline performance, future studies may benefit from adaptive or more challenging paradigms to better capture cognitive change in non-clinical populations.

#### Lack of an active control condition

The use of a waitlist control limits causal specificity, as expectancy effects, demand characteristics, or nonspecific engagement effects cannot be fully ruled out. Inclusion of an active control condition (e.g., health education, relaxation exercises, or non-meditative attention tasks) would strengthen causal inference and may also improve retention in control arms by providing participants with a structured daily activity. The absence of such a control represents an important limitation and should be addressed in future trials.

#### Constraints on crossover and washout designs

Because meditation is a self-dosed behavioral intervention with no clear biological half-life, it was not feasible to implement a traditional crossover design with a washout period. Consequently, residual or carryover effects cannot be fully disentangled from sustained intervention effects, particularly for outcomes that persisted during follow-up.

#### Sample size and statistical power

Although the study enrolled nearly 300 participants, attrition and incomplete datasets reduced the effective sample size for several analyses, particularly for physiological and cognitive outcomes. As a result, the study may have been underpowered to detect small-to-moderate effects in these domains, especially given the use of conservative multiple-comparisons corrections.

#### Generalizability

The sample consisted exclusively of undiagnosed adults, many of whom resided in or near the San Francisco Bay Area. This limits generalizability to clinical populations, individuals with limited access to technology, or populations with different cultural or socioeconomic backgrounds. Future studies should incorporate broader recruitment strategies and explicitly test SOS meditation in clinically diagnosed samples to assess translational relevance.

### Conclusions

Despite these limitations, this study demonstrates that ultra-brief, remotely delivered focused-attention meditation can meaningfully reduce internalizing symptoms in a non-clinical population. Effects were most robust for anxiety and mind wandering and were concentrated among individuals with elevated baseline symptom burden, consistent with a dimensional model of intervention responsiveness. While cognitive and physiological effects were more modest, findings highlight the feasibility of integrating low-dose meditation into scalable digital mental health frameworks. Together, these results support the potential of brief, internet-delivered meditation as a preventive strategy within digitally mediated models of care, particularly when personalized to baseline psychological profile.

## Methods

### Study Design

This study was a 16-week, fully remote randomized controlled trial with a delayed-intervention (waitlist-controlled) design (Fig. 7). The active meditation intervention lasted 8 weeks, followed by an 8-week follow-up period. The study was conducted between December 2023 and July 2025, with coordination and oversight provided by staff at Stanford University School of Medicine (California, USA).

**Fig. 6.**
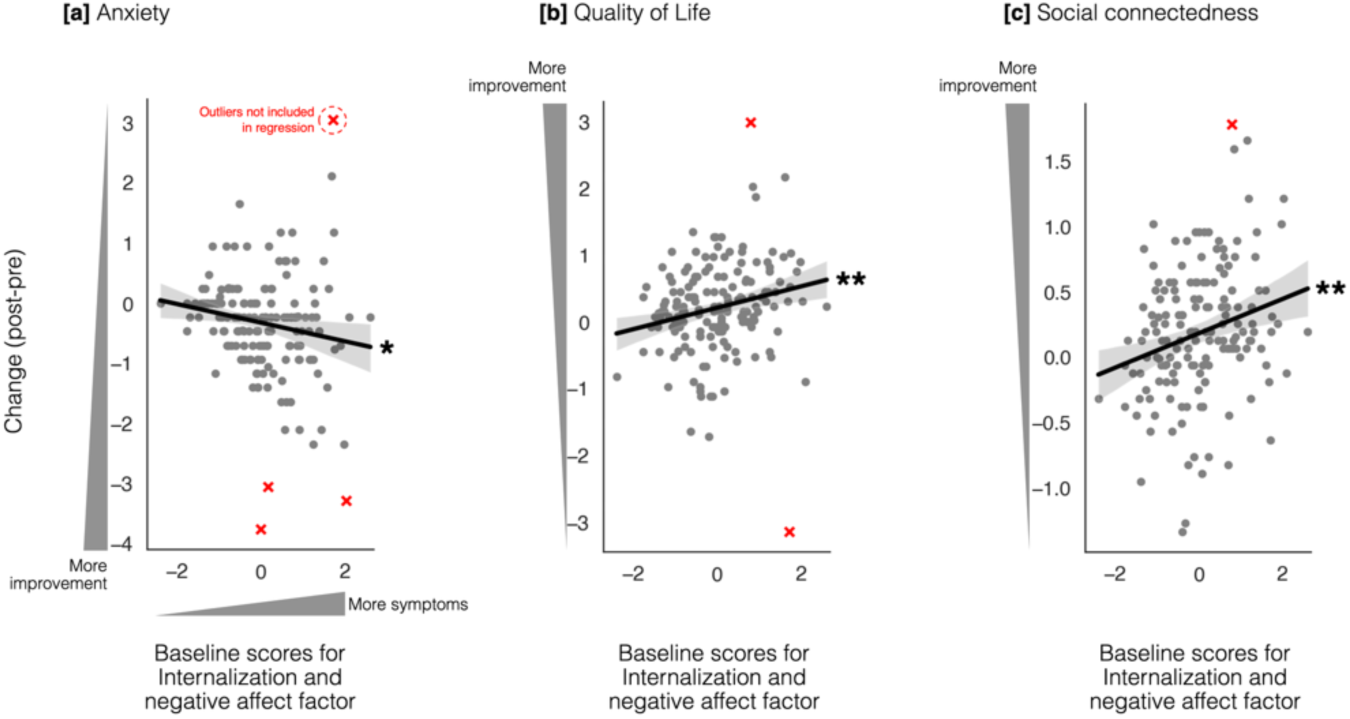
Baseline latent factors are associated with differential intervention response. (a) Partial correlation coefficients (controlling for age and sex) between baseline factor scores and post–pre (Δ) change in individual outcomes. Only associations surviving FDR correction within the domain are shown. (b–c) Representative scatterplots illustrating significant associations between baseline factor scores and Δ change in selected outcomes. Lines represent linear regression fits with 95% confidence intervals. Stars indicate FDR-corrected significance levels as in other figures.

**Fig. 7.**
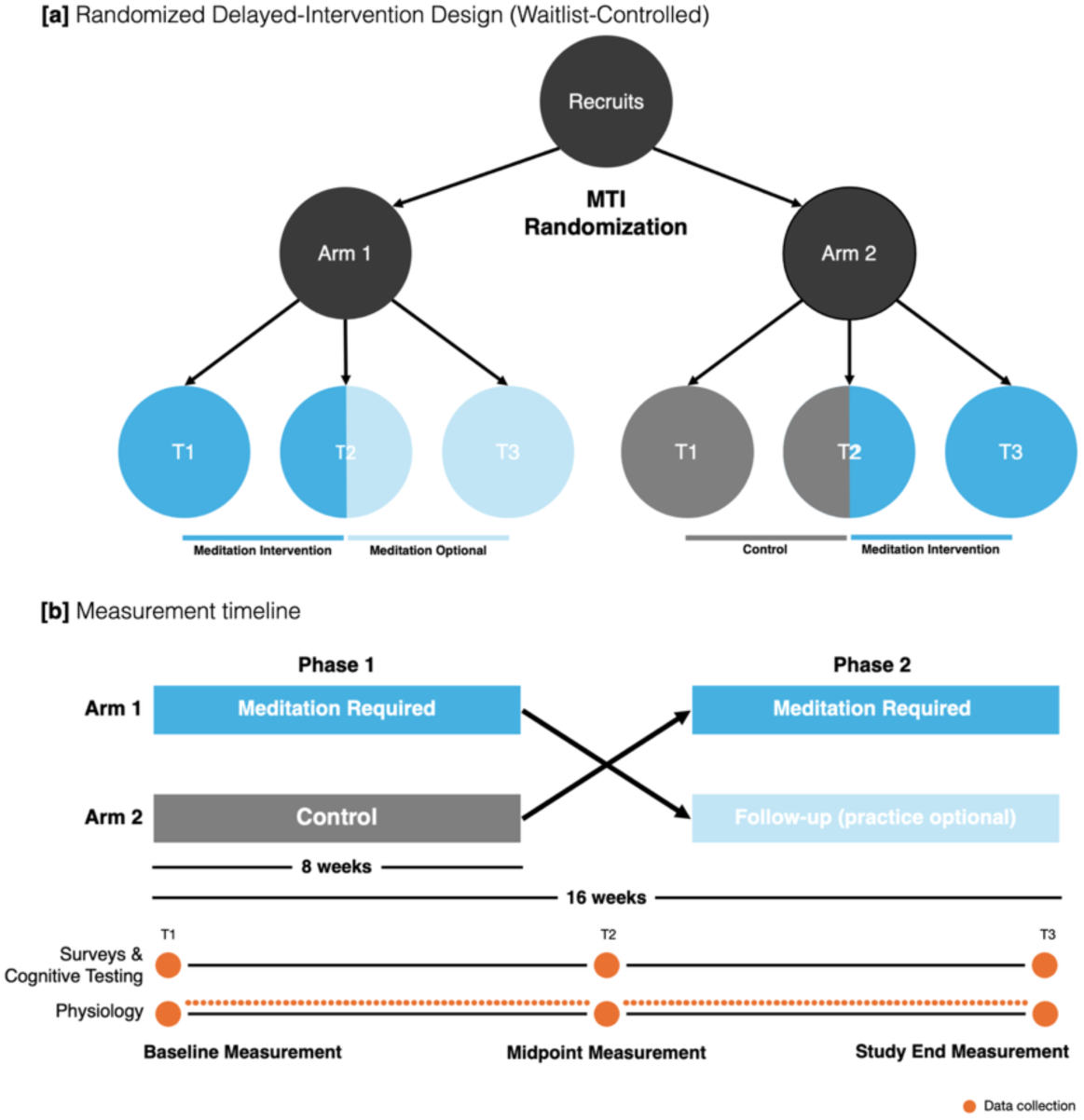
Study design and assessment timeline. (a) Participants were randomized 1:1 to either immediate intervention (Arm 1) or waitlist control with delayed intervention (Arm 2). Data were collected at three timepoints (T1: baseline; T2: week 8; T3: week 16). For Arm 1, T1–T2 corresponds to the intervention period and T2–T3 to follow-up. For Arm 2, T1–T2 corresponds to the waitlist control period and T2–T3 to the intervention period. (b) Study timeline illustrating the 16-week structure. Self-report and cognitive assessments were administered at T1, T2, and T3. Physiological measures were collected daily. No washout period was implemented.

Participants assigned to Arm 1 initiated the meditation intervention immediately following baseline assessment and completed 8 weeks of daily practice. After week 8, they entered a naturalistic follow-up phase during which continued practice was optional. Participants assigned to Arm 2 completed an initial 8-week waitlist control period prior to receiving the identical 8-week meditation intervention. This design enabled between-arm comparisons during the randomized phase (weeks 0–8) and within-participant replication in the delayed-intervention arm (weeks 8–16).

Primary self-report and cognitive outcomes were assessed at baseline (week 0), post-intervention (week 8), and follow-up (week 16). Physiological measures were collected daily throughout the study. Intervention adherence was monitored during each participant’s active meditation phase.

Because meditation is a behavioral intervention without a defined pharmacokinetic half-life, no formal washout period was implemented, and intervention and follow-up phases proceeded consecutively.

The trial was registered at ClinicalTrials.gov (Identifier: NCT06014281).

### Participant Recruitment and Eligibility

Participants were recruited across the United States between September 2023 and December 2024 via email distribution lists, social media postings, clinical trial registries, and word-of-mouth outreach. Although participation was open nationally, most enrolled participants resided in the San Francisco Bay Area.

Eligible participants were adults aged ≥18 years who expressed interest in learning meditation and were willing to complete a fully remote 16-week study. Exclusion criteria included any current or recent (within six months) psychiatric or neurological diagnosis; medical or sensory conditions that could interfere with study participation or outcome measurement (e.g., significant visual impairment including color blindness, hearing impairment, diagnosed sleep disorders such as narcolepsy, recent prolonged hospitalization); inability to wear a wrist-based fitness tracker; and prior regular meditation practice, defined as meditating ≥3 times per week for >10 minutes per session within the six months preceding enrollment.

Participants were required to reside in the United States, have reliable internet access, and be able to comply with daily data collection procedures during intervention periods. Screening was conducted online using structured questionnaires, and eligibility was confirmed by trained study staff prior to enrollment. Participants received no financial compensation beyond device provision.

All participants provided informed consent electronically prior to study participation. Consent included agreement to random assignment, repeated assessments, daily data collection during intervention phases, and secure storage of de-identified data for research purposes. The study protocol was approved by the Stanford University Institutional Review Board (IRB #68784) and registered at ClinicalTrials.gov (NCT06014281).

### Randomization

Participants were randomized in a 1:1 ratio to one of two study arms using the NIH Clinical Randomization Tool implementing the Maximum Tolerated Imbalance (MTI) procedure (Berger et al., 2003). The maximal imbalance constraint was set to three participants to maintain near-equal allocation throughout enrollment. Randomization was not stratified by age or sex.

Allocation sequences were generated prior to study initiation. Participants were assigned sequentially as they completed eligibility screening and provided informed consent, allowing balanced allocation during rolling enrollment. Study staff had no influence over allocation sequence

### Meditation Intervention

The intervention consisted of three components: (1) an initial remote training session, (2) a mid-intervention check-in session, and (3) daily self-guided meditation practice over an 8-week period.

#### Initial Training Session

Participants attended a 60-minute live remote training session conducted via secure videoconferencing. Sessions were delivered in small groups (≤12 participants) by a single instructor (author M.S.) using a standardized script to ensure consistency. When six or more participants were present, a second trained staff member (authors C.G. or S.P.) joined to maintain a maximum 6:1 participant-to-staff ratio.

The training included an overview of focused-attention meditation principles, two guided practice periods (10 mins each), and structured discussion segments following each practice period. Participants were required to keep cameras on to verify engagement and were encouraged to ask questions during the session.

#### Mid-Intervention Check-In

Approximately four weeks after initiating practice, participants attended a 30-minute remote check-in session led by trained study staff. Sessions were conducted in small groups (≤5 participants) to maintain individualized engagement. The check-in included guided practice (10 min), troubleshooting of common challenges, and reminders regarding study procedures. If scheduling conflicts arose, sessions could be rescheduled within the intervention window.

#### Daily Meditation Practice

Participants were instructed to engage in a focused-attention meditation practice for a minimum of 10 minutes per day during the 8-week intervention period (56 days). The practice was derived from a meditation approach historically associated with the organization Science of Spirituality (SOS) but was implemented in this study as a secular attentional training exercise.

To minimize expectancy effects and preserve methodological neutrality, the intervention was presented as a focused-attention meditation protocol without emphasis on spiritual framing.

Participants were provided with standardized meditation instructions (Singh, 2022), delivered verbatim during training and reproduced below to ensure intervention transparency and reproducibility:

1. *Close your eyes gently and relax, as you would when preparing to sleep*.
2. *Keep your attention alert and avoid straining your eyes, crossing them, or directing them upward*.
3. *Direct your inner gaze approximately 8–10 inches into the field of darkness in front of you along the horizontal plane*.
4. *Silently repeat any calming word*.
5. *Remain still and observe internal experience calmly, as though watching images arise on a screen*.

Participants were permitted flexibility in accumulating their daily practice time (e.g., one 10-minute session or multiple shorter sessions (≥5 minutes) for a total of ≥10 minutes every day. Continued meditation beyond the required 8-week period was optional during follow-up phases.

### Outcome Measures

Outcomes were assessed across three primary domains: psychological self-report, cognitive performance, and physiological regulation.

#### Primary Outcomes

Primary psychological outcomes included generalized anxiety (GAD-7), sleep disturbance (Pittsburgh Sleep Quality Index), and mind wandering (Mind Wandering Questionnaire). These instruments are validated measures widely used in clinical and non-clinical populations (Spitzer et al., 2006; Buysse et al., 1989; Mrazek et al., 2013).

Primary physiological outcomes included resting heart rate (RHR) and heart rate variability (HRV), derived from nightly recordings using Fitbit Charge 6 wearable devices. Daily measurements were aggregated within predefined study windows for analysis.

Primary cognitive outcomes included performance on web-based Stroop and 2-back tasks administered via the Gorilla.sc platform. From these tasks, accuracy and reaction time metrics were derived to index executive function and working memory.

#### Secondary Outcomes

Secondary psychological measures included perceived stress (PSS), depressive symptoms (PHQ-8), quality of life, social connectedness, rumination and reflection, and character strengths. Secondary physiological outcomes included wearable-derived sleep quality metrics.

A detailed summary of all primary and secondary measures, including assessment timing and operational definitions, is provided in Tables 3 and 4.

**Table 3:**
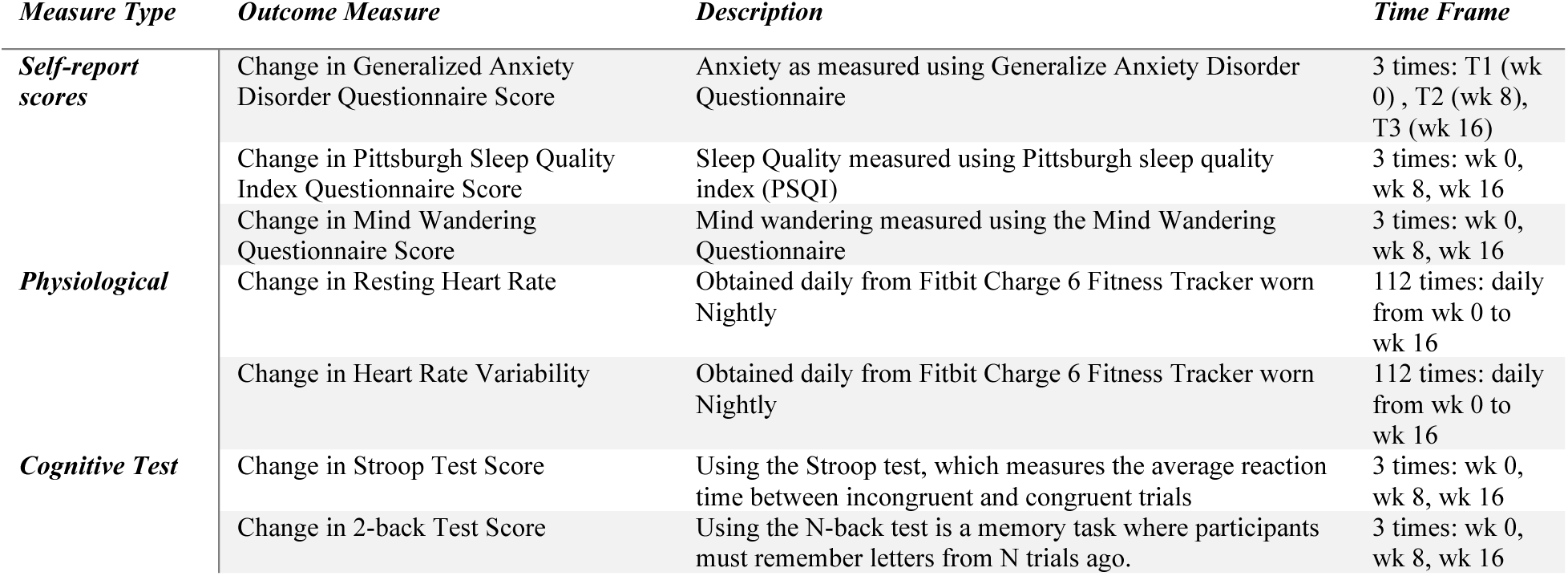
Primary Measures. A summary table listing primary measures of interest, their description, and timepoints at which the data were collected.

| *Measure Type* | *Outcome Measure* | *Description* | *Time Frame* |
| --- | --- | --- | --- |
| **Self-report scores** | Change in Generalized Anxiety Disorder Questionnaire Score | Anxiety as measured using Generalize Anxiety Disorder Questionnaire | 3 times: T1 (wk 0), T2 (wk 8), T3 (wk 16) |
|  | Change in Pittsburgh Sleep Quality Index Questionnaire Score | Sleep Quality measured using Pittsburgh sleep quality index (PSQI) | 3 times: wk 0, wk 8, wk 16 |
|  | Change in Mind Wandering Questionnaire Score | Mind wandering measured using the Mind Wandering Questionnaire | 3 times: wk 0, wk 8, wk 16 |
| **Physiological** | Change in Resting Heart Rate | Obtained daily from Fitbit Charge 6 Fitness Tracker worn Nightly | 112 times: daily from wk 0 to wk 16 |
|  | Change in Heart Rate Variability | Obtained daily from Fitbit Charge 6 Fitness Tracker worn Nightly | 112 times: daily from wk 0 to wk 16 |
| **Cognitive Test** | Change in Stroop Test Score | Using the Stroop test, which measures the average reaction time between incongruent and congruent trials | 3 times: wk 0, wk 8, wk 16 |
|  | Change in 2-back Test Score | Using the N-back test is a memory task where participants must remember letters from N trials ago. | 3 times: wk 0, wk 8, wk 16 |

**Table 4:** Secondary Measures. A summary table listing secondary measures of interest, their description, and timepoints at which the data were collected.

| *Measure Type* | *Outcome Measure* | *Description* | *Time Frame* |
| --- | --- | --- | --- |
| **Self-report scores** | Change in Stress Scale Measure Questionnaire Score | Stress measured using Perceived Stress Scale measure | 3 times: T1 (wk 0), T2 (wk 8), T3 (wk 16) |
|  | Change in Depression Questionnaire Score | Depression measured using Patient Health Questionnaire | 3 times: wk 0, wk 8, wk 16 |
|  | Change in Quality of Life Scale Measures Questionnaire Score | Quality of life measured using Quality of Life Scale measures | 3 times: wk 0, wk 8, wk 16 |
|  | Change Social Connectedness Scale-Revised Questionnaire Score | Social Connectedness measured using Social Connectedness Scale-Revised | 3 times: wk 0, wk 8, wk 16 |
|  | Change in Rumination Reflection Questionnaire Score | Rumination and reflection scales measured using Rumination Reflection Questionnaire | 3 times: wk 0, wk 8, wk 16 |
|  | Change in Brief Strengths Scale Questionnaire Score | Strengths scale measured using Brief Strengths Scale measures individuals' Temperance Strength, Intellectual Strength, and Interpersonal Strength. | 3 times: wk 0, wk 8, wk 16 |
| **Physiological** | Change in Sleep Quality Measure | Obtained daily from Fitbit Charge 6 Fitness Tracker worn Nightly | 112 times: daily from wk 0 to wk 16 |

All outcome measures, study design, etc. were preregistered before data collection (ClinicalTrials.gov: NCT06014281).

### Data Collection

All study data were collected remotely using secure, web-based platforms. Participants were provided with written instructions and contact information for study staff to address technical or procedural questions. Because several questionnaires assessed sensitive mental health domains, participants were permitted to skip individual items if desired.

#### Self-Report Measures

Self-report data were collected using REDCap, a secure web-based data capture system. Demographic information was obtained at baseline to assess eligibility and characterize the sample. These data included age, sex, ethnicity, education level, employment status, marital status, and prior meditation experience and were not reassessed at follow-up.

Validated mental health and well-being questionnaires were administered at baseline (T1), week 8 (T2), and week 16 (T3). Measures included the Generalized Anxiety Disorder–7 (GAD-7), Social Connectedness Scale–Revised (SCS-R), Pittsburgh Sleep Quality Index (PSQI), Rumination–Reflection Questionnaire (RRQ), Quality of Life Scale (QOLS), Patient Health Questionnaire–8 (PHQ-8), Perceived Stress Scale (PSS), Brief Strengths Scale–12 (BSS-12), and Mind Wandering Questionnaire (MWQ).

Responses were reviewed upon submission. Participants whose scores fell within clinically severe ranges were contacted by trained study staff and provided with appropriate mental health resources according to a predefined safety protocol.

#### Meditation Adherence Monitoring

During each participant’s 8-week intervention phase (Arm 1: T1–T2; Arm 2: T2–T3), daily meditation logs were completed via REDCap. Participants recorded duration of practice, subjective focus, subjective calm following practice, and date of completion. Participants reporting fewer than four days of meditation within a given week were contacted to support adherence. Logs submitted outside the required 8-week intervention window were not included in analyses.

#### Physiological Data Collection

Participants were provided with a Fitbit Charge 6 wearable device prior to study initiation. Devices were worn nightly from approximately 10–15 minutes before sleep until 10–15 minutes after waking, yielding up to 112 nights of data across the 16-week study. Although participants could wear the device during daytime hours, only nocturnal data were analyzed.

Physiological data were aggregated using Fitabase, a third-party platform for wearable data extraction. Resting heart rate (RHR) and heart rate variability (HRV) were extracted for analysis. Sleep-stage metrics were collected but are not reported in the present manuscript.

#### Cognitive Task Administration

Cognitive performance was assessed at T1, T2, and T3 using web-based Stroop and 2-back tasks delivered via the Gorilla.sc platform. Each task was administered in three runs per timepoint (2-back: 30 trials per run; Stroop: 27 trials per run). Across the full study, participants completed up to nine runs per task.

To reduce variability associated with remote testing environments, occasional misunderstanding of task instructions, or incomplete runs, the run with the highest accuracy at each timepoint was selected for analysis. Task and run order were randomized across participants and sessions.

The Stroop task measured selective attention and interference control through accuracy and reaction time differences between congruent and incongruent color-word trials. The 2-back task assessed working memory by requiring participants to identify matches between the current stimulus and the letter presented two trials earlier. Accuracy and reaction time were recorded for all trials.

### Data Analysis

All analyses were conducted in accordance with the preregistered study design. Analyses were performed using R (v4.2.0) and Python (v3.14.0). All tests were two-sided unless otherwise specified. False discovery rate (FDR; Benjamini–Hochberg) correction was applied within predefined outcome families where indicated.

#### Handling of Missing Data

Missing data were addressed using multiple imputation by chained equations (MICE; van Buuren & Groothuis-Oudshoorn, 2011)) with predictive mean matching. Imputation was performed separately within each outcome domain and analytic window to preserve the temporal structure of the study. Fifty imputed datasets were generated using 20 iterations per dataset. Study arm and sex were not imputed; age was imputed when missing.

For between-arm comparisons at week 8 (T1–T2), imputation models included baseline (T1) and post-intervention (T2) scores, study arm, sex, and age. For within-arm analyses (replication in Arm 2 and follow-up in Arm 1; T2–T3), imputation was restricted to the T2–T3 window, with week 8 and week 16 scores estimated, and baseline score, sex, and age included as auxiliary variables.

To preserve the randomized structure for Arm 2 at week 8, a post-imputation last-observation-carried-forward (LOCF) rule was applied when week-8 values were missing but baseline values were observed. Estimates were pooled across imputations using Rubin’s rules.

Analytic estimates (effects, standard errors, confidence intervals, and p-values) were pooled across imputations using Rubin’s rules (Rubin, 1987), as implemented in the mice package.

#### Primary Analytic Framework

For all outcome domains, we implemented a three-part analytic strategy:

1. Discovery (between-group) analysis comparing Arm 1 (intervention) vs Arm 2 (control) during the randomized phase (T1–T2)
2. Replication (within-group) analysis in Arm 2 during delayed intervention (T2–T3)
3. Sustainability analysis in Arm 1 during follow-up (T2–T3)

Age and sex were included as covariates in all adjusted between-group models.

#### Self-Report Outcomes

Between-group effects at week 8 were tested using ANCOVA:

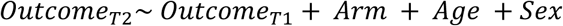

Within-arm changes (replication and follow-up) were assessed using paired-sample t-tests.

To evaluate differential effects by baseline severity, pre–post changes were pooled across intervention periods (Arm 1: T1–T2; Arm 2: T2–T3). Participants were grouped by established clinical cutoffs (when available) or quartiles. Paired t-tests were conducted within subgroups, with FDR correction applied across subgroup comparisons.

#### Physiological Outcomes

Primary physiological outcomes included resting heart rate (RHR) and heart rate variability (HRV). Between-group comparisons at week 8 were analyzed using ANCOVA models analogous to those used for self-report outcomes. Within-arm replication and follow-up analyses were conducted using paired t-tests.

Baseline-stratified analyses were performed using pooled intervention windows and quartile grouping, with FDR correction applied within physiological outcomes.

#### Cognitive Outcomes

Cognitive outcomes were derived from Stroop and 2-back tasks. At each timepoint, the run with the highest accuracy was selected for analysis to reduce variability associated with remote testing conditions.

Primary cognitive metrics included accuracy and reaction time for Stroop congruent and incongruent trials and total accuracy and reaction time for 2-back trials. Analyses followed the same discovery, replication, and follow-up framework described above, using ANCOVA for between-group comparisons and paired t-tests for within-group analyses. Baseline-stratified analyses used quartile grouping due to the absence of clinical cutoffs.

#### Exploratory Factor Analysis

Exploratory factor analysis (EFA) was conducted on baseline self-report measures. Factorability was assessed using Bartlett’s test of sphericity and the Kaiser–Meyer–Olkin statistic. Horn’s parallel analysis determined the number of factors retained (factor-analyzer 0.5.1; scikit-learn 1.1.1; scipy 1.13.1; statsmodels 0.14.4).

A two-factor oblimin-rotated solution was estimated. Loadings ≥ |0.40| were retained for interpretability. Factor scores were extracted from the baseline-fitted model (Stevens, 1992).

Between-group comparisons at week 8 were analyzed using ANCOVA models analogous to those used for self-report outcomes. Within-arm replication and follow-up analyses were conducted using paired t-tests.

#### Associations Between Baseline Factors and Change

To examine individual differences in responsiveness, partial correlations were computed between baseline factor scores and standardized pre–post change scores within self-report, physiological, and cognitive domains, controlling for age and sex. FDR correction was applied within each outcome domain. Only associations surviving FDR correction were reported.

## Data availability

De-identified data is available upon reasonable request from the corresponding author. Code is available at the *Brain Dynamics Lab GitHub* page.

## Acknowledgements

We would like to thank the participants. We also thank the philanthropic funding from the IMIH (Chicago) and FFCA (San Francisco) to M.S.

## Author information

**C.G.** Conceptualization, Methodology, Validation, Software, Formal Analysis, Investigation, Data Curation, Writing – Original Draft, Visualization, Project Administration

**S.P.** Conceptualization, Methodology, Validation, Software, Formal Analysis, Investigation, Data Curation, Writing – Original Draft, Visualization

**S.Q.** Conceptualization, Writing - Review & Editing

**Sa. F.** Investigation, Writing – Original Draft

**I.E.** Recruiting and data entry

**S.M.** Data gathering and preprocessing

**N.B.** Developing Fitbit data acquisition platform

**Su. F.** Data entry and preprocessing

**R.B.** Conceptualization, Writing - Review & Editing

**D.S.** Conceptualization, Writing - Review & Editing, Supervision

**M.S**. Conceptualization, Methodology, Investigation, Writing – Original Draft, Project Administration, Supervision, Funding acquisition

## Ethics declaration

This study was conducted according to the ethical guidelines outlined in the *Declaration of Helsinki*. Approval was obtained, and the study was monitored by the Stanford Institutional Review Board. All participants provided informed consent and were informed of the option to withdraw from the study at any time without penalty. There are no financial interests to report.

