## Supplementary Information for "A fully remote randomized controlled trial of an ultra-brief digital meditation intervention reduces internalizing symptoms"

### Supplemental Tables

**Table S1: Primary psychological outcomes – imputed between-arm ANCOVA at $T_2$ (week 8)**

| *Measure* | *Arm 1 n* | *Arm 2 n* | *Adj. Mean ± SE (Arm 1)* | *Adj. Mean ± SE (Arm 2)* | *F* | *p (raw)* | *p (FDR-BH)* | *Partial η²* |
|---|---|---|---|---|---|---|---|---|
| ***GAD-7*** | 121 | 114 | 4.04 ± 0.34 | 5.07 ± 0.33 | 4.989 | **0.0255** | **0.0383** | 0.001 |
| ***PSQI*** | 120 | 114 | 6.94 ± 0.22 | 7.37 ± 0.21 | 2.045 | 0.1528 | 0.1528 | 0 |
| ***MWQ*** | 120 | 113 | 15.52 ± 0.37 | 17.11 ± 0.37 | 9.817 | **0.0017** | **0.0052** | 0.002 |

**Table S2: Primary psychological outcomes – imputed replication analysis - arm 2 (within-subject) at $T_3$ vs $T_2$ (week 16 vs week 8) t-test**

| *Measure* | *n* | *$T_2$ Mean ± SEM* | *$T_3$ Mean ± SEM* | *Δ ($T_3$–$T_2$)* | *95% CI Low* | *95% CI High* | *t* | *df* | *p (raw)* | *p (FDR-BH)* | *Cohen $d_z$* |
|---|---|---|---|---|---|---|---|---|---|---|---|
| ***GAD-7*** | 114 | 5.04 ± 0.39 | 3.74 ± 0.36 | -1.29 | -1.94 | -0.63 | -3.849 | 3526.706 | **0.0001** | **0.0002** | -0.36 |
| ***PSQI*** | 114 | 7.45 ± 0.24 | 7.21 ± 0.24 | -0.25 | -0.7 | 0.21 | -1.057 | 3118.112 | 0.2906 | 0.2906 | -0.1 |
| ***MWQ*** | 113 | 17.03 ± 0.48 | 14.88 ± 0.51 | -2.06 | -3.03 | -1.08 | -4.146 | 2405.242 | **0** | **0.0001** | -0.39 |

**Table S3: Primary psychological outcomes – imputed sustainability analysis - arm 1 (within-subject) at $T_3$ vs $T_2$ (week 16 vs week 8) t-test**

| *Measure* | *n* | *Week-8 Mean ± SEM* | *Week-16 Mean ± SEM* | *Δ ($T_3$-$T_2$)* | *95% CI Low* | *95% CI High* | *t* | *df* | *p (raw)* | *p (FDR-BH)* | *Cohen $d_z$* |
|---|---|---|---|---|---|---|---|---|---|---|---|
| ***GAD-7*** | 103 | 4.25 ± 0.40 | 3.83 ± 0.37 | -0.38 | -1.22 | 0.46 | -0.881 | 1812.6 | 0.3785 | 0.754 | -0.09 |
| ***PSQI*** | 100 | 7.01 ± 0.21 | 7.15 ± 0.22 | 0.11 | -0.34 | 0.55 | 0.47 | 4700.448 | 0.6385 | 0.754 | 0.05 |
| ***MWQ*** | 98 | 15.53 ± 0.51 | 15.32 ± 0.50 | -0.09 | -0.67 | 0.49 | -0.313 | 1835.62 | 0.754 | 0.754 | -0.03 |

**Table S4: Primary psychological outcomes – imputed clinical-impact analysis – arm 1 and arm 2 pooled over intervention period and stratified by clinical severity - post vs pre t-test**

| *Measure* | *Quartile/ Category* | *n* | *Pre mean* | *Pre SEM* | *Post mean* | *Post SEM* | *Δ (Post-Pre)* | *CI low* | *CI High* | *t* | *df* | *p (raw)* | *p (FDR-BH)* |
|---|---|---|---|---|---|---|---|---|---|---|---|---|---|
| ***GAD-7*** | Minimal (0–4) | 106 | 1.84 | 0.14 | 2.04 | 0.25 | 0.2 | -0.24 | 0.64 | 0.904 | 2862.1 | 0.3659 | 0.4878 |
| ***GAD-7*** | Mild (5–9) | 89 | 6.84 | 0.15 | 5 | 0.42 | -1.84 | -2.63 | -1.05 | -4.562 | 1979.8 | **0** | **0** |
| ***GAD-7*** | Moderate (10–14) | 22 | 11.95 | 0.28 | 7.48 | 0.81 | -4.48 | -6.06 | -2.89 | -5.529 | 2739.5 | **0** | **0** |
| ***GAD-7*** | Severe (15–21) | 16 | 17.38 | 0.46 | 8.33 | 1.56 | -9.05 | -12.32 | -5.78 | -5.429 | 745.2 | **0** | **0** |
| ***PSQI*** | Q1 (<25th) | 32 | 3.78 | 0.33 | 5 | 0.43 | 1.22 | 0.13 | 2.31 | 2.195 | 3788.5 | **0.0282** | **0.0424** |
| ***PSQI*** | Q2 (25–50th) | 66 | 6.52 | 0.06 | 6.68 | 0.19 | 0.17 | -0.23 | 0.57 | 0.822 | 1757.9 | 0.4114 | 0.4936 |

| | | | | | | | | | | | | | |
|---|---|---|---|---|---|---|---|---|---|---|---|---|---|
| ***PSQI*** | Q3 (50–75th) | 47 | 8 | 0 | 7.24 | 0.28 | -0.76 | -1.32 | -0.21 | -2.723 | 739.7 | **0.0066** | **0.0114** |
| ***PSQI*** | Q4 (≥75th) | 87 | 10.25 | 0.15 | 8.23 | 0.29 | -2.02 | -2.59 | -1.45 | -6.944 | 1522.5 | **0** | **0** |
| ***MWQ*** | Q1 (<25th) | 58 | 10.52 | 0.31 | 10.62 | 0.52 | 0.1 | -0.85 | 1.05 | 0.211 | 2179.7 | 0.8328 | 0.9085 |
| ***MWQ*** | Q2 (25–50th) | 50 | 15.12 | 0.12 | 15.12 | 0.58 | 0 | -1.13 | 1.14 | 0.004 | 740.4 | 0.9967 | 0.9967 |
| ***MWQ*** | Q3 (50–75th) | 64 | 18.66 | 0.15 | 16.6 | 0.52 | -2.06 | -3.06 | -1.05 | -4.014 | 1533 | **0.0001** | **0.0001** |
| ***MWQ*** | Q4 (≥75th) | 59 | 23.85 | 0.33 | 19.1 | 0.72 | -4.75 | -6.21 | -3.29 | -6.363 | 1676.7 | **0** | **0** |

**Table S5: Secondary psychological outcomes – imputed between-arm ANCOVA at $T_2$ (week 8)**

| *Measure* | *Arm 1 n* | *Arm 2 n* | *Adj. Mean ± SE (Arm 1)* | *Adj. Mean ± SE (Arm 2)* | *F* | *p (raw)* | *p (FDR-BH)* | *Partial η²* |
|---|---|---|---|---|---|---|---|---|
| ***SCS-R*** | 120 | 114 | 88.95 ± 0.87 | 86.43 ± 0.82 | 4.618 | **0.0317** | 0.0952 | 0.002 |
| ***RRQ-Rumination*** | 120 | 114 | 3.04 ± 0.05 | 3.21 ± 0.05 | 6.55 | **0.0106** | 0.095 | 0.003 |
| ***PSS*** | 120 | 114 | 13.61 ± 0.48 | 14.84 ± 0.45 | 3.649 | 0.0562 | 0.1264 | 0.001 |
| ***QOLS*** | 112 | 105 | 82.89 ± 0.95 | 80.11 ± 0.87 | 4.705 | **0.0303** | 0.0952 | 0.004 |
| ***PHQ-8*** | 120 | 114 | 3.30 ± 0.28 | 3.72 ± 0.28 | 1.206 | 0.2721 | 0.4082 | 0 |
| ***RRQ-Reflection*** | 120 | 114 | 3.64 ± 0.04 | 3.56 ± 0.04 | 1.468 | 0.2258 | 0.4064 | 0 |
| ***BSS-12-Interpersonal*** | 121 | 115 | 6.17 ± 0.04 | 6.12 ± 0.04 | 0.748 | 0.3872 | 0.4979 | 0 |
| ***BSS-12-Temperance*** | 121 | 115 | 5.65 ± 0.05 | 5.69 ± 0.05 | 0.211 | 0.6462 | 0.727 | 0 |
| ***BSS-12-Intellectual*** | 121 | 115 | 6.04 ± 0.05 | 6.03 ± 0.05 | 0.033 | 0.8569 | 0.8569 | 0 |

**Table S6: Secondary psychological outcomes – imputed replication analysis - arm 2 (within-subject) at $T_3$ vs $T_2$ (week 16 vs week 8) t-test**

| *Measure* | *n* | *$T_2$ Mean ± SEM* | *$T_3$ Mean ± SEM* | *Δ ($T_3$−$T_2$)* | *95% CI Low* | *95% CI High* | *t* | *df* | *p (raw)* | *p (FDR-BH)* | *Cohen $d_z$* |
|---|---|---|---|---|---|---|---|---|---|---|---|
| ***SCS-R*** | 114 | 85.56 ± 1.41 | 88.97 ± 1.48 | 3.38 | 1.6 | 5.16 | 3.729 | 887.743 | **0.0002** | **0.0006** | 0.35 |
| ***RRQ-Rumination*** | 114 | 3.24 ± 0.08 | 3.00 ± 0.09 | -0.26 | -0.37 | -0.14 | -4.199 | 880.2194 | **0** | **0.0003** | -0.39 |
| ***PSS*** | 114 | 15.13 ± 0.58 | 13.69 ± 0.65 | -1.39 | -2.47 | -0.3 | -2.496 | 2809.0068 | **0.0126** | **0.0227** | -0.23 |
| ***QOLS*** | 109 | 78.95 ± 1.24 | 83.32 ± 1.36 | 3.71 | 1.75 | 5.67 | 3.716 | 871.5425 | **0.0002** | **0.0006** | 0.36 |
| ***PHQ-8*** | 114 | 3.77 ± 0.31 | 3.57 ± 0.39 | -0.21 | -0.83 | 0.41 | -0.665 | 1927.9085 | 0.506 | 0.5692 | -0.06 |
| ***RRQ-Reflection*** | 114 | 3.56 ± 0.07 | 3.66 ± 0.07 | 0.1 | 0.03 | 0.18 | 2.633 | 828.9516 | **0.0086** | **0.0194** | 0.25 |
| ***BSS-12-Interpersonal*** | 115 | 6.11 ± 0.06 | 6.01 ± 0.10 | -0.1 | -0.28 | 0.07 | -1.17 | 790.2727 | 0.2422 | 0.3633 | -0.11 |
| ***BSS-12-Temperance*** | 115 | 5.68 ± 0.08 | 5.71 ± 0.10 | 0.03 | -0.14 | 0.19 | 0.332 | 636.4889 | 0.7402 | 0.7402 | 0.03 |

| | | | | | | | | | | | |
|---|---|---|---|---|---|---|---|---|---|---|---|
| ***BSS-12-Intellectual*** | 115 | 24.03 ± 0.25 | 23.69 ± 0.38 | -0.34 | -1.04 | 0.36 | -0.959 | 694.7354 | 0.3379 | 0.4344 | -0.09 |

**Table S7: Secondary psychological outcomes – imputed sustainability analysis - arm 1 (within-subject) at $T_3$ vs $T_2$ (week 16 vs week 8) t-test**

| *Measure* | *n* | *$T_2$ Mean ± SEM* | *$T_3$ Mean ± SEM* | *Δ ($T_3$-$T_2$)* | *95% CI Low* | *95% CI High* | *t* | *df* | *p (raw)* | *p (FDR-BH)* | *Cohen $d_z$* |
|---|---|---|---|---|---|---|---|---|---|---|---|
| ***SCS-R*** | 101 | 88.83 ± 1.43 | 89.58 ± 1.54 | 0.42 | -1.18 | 2.01 | 0.513 | 1597.228 | 0.6082 | 0.6622 | 0.05 |
| ***RRQ-Rumination*** | 100 | 3.04 ± 0.08 | 2.99 ± 0.09 | -0.04 | -0.14 | 0.05 | -0.89 | 2637.342 | 0.3737 | 0.6622 | -0.09 |
| ***PSS*** | 103 | 13.76 ± 0.70 | 14.20 ± 0.72 | 0.49 | -0.8 | 1.77 | 0.746 | 1449.469 | 0.4558 | 0.6622 | 0.07 |
| ***QOLS*** | 92 | 83.53 ± 1.42 | 83.57 ± 1.36 | -0.54 | -2.94 | 1.87 | -0.437 | 1456.559 | 0.6622 | 0.6622 | -0.05 |
| ***PHQ-8*** | 103 | 3.52 ± 0.38 | 3.18 ± 0.37 | -0.36 | -1.1 | 0.38 | -0.953 | 2955.191 | 0.3409 | 0.6622 | -0.09 |
| ***RRQ-Reflection*** | 100 | 3.64 ± 0.07 | 3.66 ± 0.08 | 0.03 | -0.05 | 0.1 | 0.646 | 1108.355 | 0.5186 | 0.6622 | 0.06 |
| ***BSS-12-Interpersonal*** | 102 | 6.19 ± 0.07 | 6.25 ± 0.07 | 0.06 | -0.04 | 0.16 | 1.149 | 1203.281 | 0.2507 | 0.6622 | 0.11 |
| ***BSS-12-Temperance*** | 102 | 5.63 ± 0.09 | 5.76 ± 0.08 | 0.14 | 0.03 | 0.24 | 2.606 | 1324.756 | **0.0093** | 0.0555 | 0.26 |
| ***BSS-12-Intellectual*** | 102 | 6.01 ± 0.07 | 6.16 ± 0.07 | 0.14 | 0.03 | 0.26 | 2.506 | 1333.264 | **0.0123** | 0.0555 | 0.25 |

**Table S8: Secondary psychological outcomes – imputed clinical-impact analysis – arm 1 and arm 2 pooled over intervention period and stratified by clinical severity - post vs pre t-test**

| *Measure* | *Quartile/ Category* | *n* | *Pre mean* | *Pre SEM* | *Post mean* | *Post SEM* | *Δ (Post-Pre)* | *CI low* | *CI High* | *t* | *df* | *p (raw)* | *p (FDR-BH)* |
|---|---|---|---|---|---|---|---|---|---|---|---|---|---|
| ***SCS-R*** | Q1 (<25th) | 58 | 66.41 | 1.08 | 73.31 | 1.59 | 6.89 | 4.78 | 9.01 | 6.402 | 637.8 | **0** | **0** |
| ***SCS-R*** | Q2 (25–50th) | 57 | 82.04 | 0.38 | 86.13 | 1.58 | 4.1 | 0.98 | 7.22 | 2.581 | 694.3 | **0.01** | **0.0258** |
| ***SCS-R*** | Q3 (50–75th) | 58 | 92.86 | 0.39 | 91.76 | 1.28 | -1.11 | -3.51 | 1.3 | -0.903 | 2126.1 | 0.3668 | 0.4248 |
| ***SCS-R*** | Q4 (≥75th) | 59 | 105.03 | 0.6 | 103.73 | 0.99 | -1.3 | -3.16 | 0.55 | -1.379 | 4465.9 | 0.1678 | 0.2251 |
| ***RRQ-Rumination*** | Q1 (<25th) | 58 | 2.12 | 0.06 | 2.06 | 0.07 | -0.06 | -0.19 | 0.07 | -0.892 | 1712.4 | 0.3724 | 0.4248 |
| ***RRQ-Rumination*** | Q2 (25–50th) | 58 | 3.14 | 0.03 | 2.95 | 0.08 | -0.19 | -0.34 | -0.03 | -2.361 | 915.1 | **0.0184** | **0.0412** |
| ***RRQ-Rumination*** | Q3 (50–75th) | 51 | 3.68 | 0.02 | 3.31 | 0.09 | -0.36 | -0.53 | -0.2 | -4.265 | 926.5 | **0** | **0.0002** |
| ***RRQ-Rumination*** | Q4 (≥75th) | 65 | 4.23 | 0.03 | 3.77 | 0.08 | -0.46 | -0.61 | -0.31 | -6.085 | 2504.1 | **0** | **0** |
| ***PSS*** | Low (0, 13) | 84 | 9 | 0.32 | 9.26 | 0.6 | 0.26 | -0.82 | 1.35 | 0.477 | 4340.4 | 0.6336 | 0.6517 |
| ***PSS*** | Moderate (14, 27) | 138 | 18.99 | 0.33 | 16.09 | 0.52 | -2.9 | -3.84 | -1.97 | -6.093 | 1535 | **0** | **0** |
| ***PSS*** | High (28, 40) | 10 | 30.2 | 0.59 | 23.74 | 2.45 | -6.46 | -10.97 | -1.95 | -2.808 | 2355.2 | **0.005** | **0.0151** |
| ***QOLS*** | Q1 (<25th) | 49 | 62.37 | 0.81 | 70.87 | 1.66 | 8.5 | 5.37 | 11.63 | 5.335 | 714.4 | **0** | **0** |
| ***QOLS*** | Q2 (25–50th) | 57 | 75.91 | 0.48 | 79.77 | 1.4 | 3.86 | 1.22 | 6.5 | 2.869 | 706.7 | **0.0042** | **0.0139** |

| | | | | | | | | | | | | | |
|---|---|---|---|---|---|---|---|---|---|---|---|---|---|
| ***QOLS*** | Q3 (50–75th) | 50 | 86.52 | 0.43 | 85.17 | 1.55 | -1.35 | -4.35 | 1.65 | -0.882 | 1987.6 | 0.3776 | 0.4248 |
| ***QOLS*** | Q4 (≥75th) | 57 | 96.97 | 0.69 | 95.43 | 1.26 | -1.54 | -3.89 | 0.81 | -1.288 | 1372.4 | 0.198 | 0.2458 |
| ***PHQ-8*** | Minimal (<5) | 143 | 1.76 | 0.12 | 2.1 | 0.2 | 0.35 | -0.03 | 0.72 | 1.815 | 2143.1 | 0.0696 | 0.1109 |
| ***PHQ-8*** | Mild (5, 9) | 63 | 6.79 | 0.16 | 4.76 | 0.49 | -2.04 | -3.05 | -1.02 | -3.945 | 1648.5 | **0.0001** | **0.0004** |
| ***PHQ-8*** | Moderate (10, 14) | 11 | 11.36 | 0.39 | 7.63 | 1.43 | -3.73 | -6.61 | -0.85 | -2.54 | 1307.9 | **0.0112** | **0.0269** |
| ***PHQ-8*** | Moderately severe (15, 19) | 12 | 16.75 | 0.45 | 9.36 | 2.22 | -7.39 | -11.44 | -3.34 | -3.577 | 1082.9 | **0.0004** | **0.0015** |
| ***PHQ-8*** | Severe (>20) | 3 | 22.67 | 1.33 | 13.44 | 5.21 | -9.23 | -21.48 | 3.03 | -1.477 | 2587.2 | 0.1399 | 0.2014 |
| ***RRQ-Reflection*** | Q1 (<25th) | 56 | 2.63 | 0.05 | 2.96 | 0.09 | 0.34 | 0.17 | 0.5 | 4.069 | 4128.1 | **0** | **0.0003** |
| ***RRQ-Reflection*** | Q2 (25–50th) | 49 | 3.3 | 0.02 | 3.42 | 0.07 | 0.12 | 0 | 0.25 | 1.928 | 2379 | 0.054 | 0.0925 |
| ***RRQ-Reflection*** | Q3 (50–75th) | 69 | 3.76 | 0.02 | 3.76 | 0.06 | 0.01 | -0.1 | 0.11 | 0.132 | 780.8 | 0.8949 | 0.8949 |
| ***RRQ-Reflection*** | Q4 (≥75th) | 58 | 4.44 | 0.04 | 4.35 | 0.06 | -0.09 | -0.19 | 0.01 | -1.807 | 4007.2 | 0.0708 | 0.1109 |
| ***BSS-12-Interpersonal*** | Q1 (<25th) | 47 | 5.14 | 0.06 | 5.37 | 0.12 | 0.23 | 0.02 | 0.44 | 2.113 | 1681.7 | **0.0347** | 0.0658 |
| ***BSS-12-Interpersonal*** | Q2 (25–50th) | 59 | 5.91 | 0.02 | 5.95 | 0.07 | 0.04 | -0.09 | 0.17 | 0.619 | 842.6 | 0.5359 | 0.5675 |
| ***BSS-12-Interpersonal*** | Q3 (50–75th) | 54 | 6.38 | 0.02 | 6.32 | 0.07 | -0.05 | -0.2 | 0.1 | -0.672 | 1383.9 | 0.5014 | 0.547 |
| ***BSS-12-Interpersonal*** | Q4 (≥75th) | 74 | 6.88 | 0.01 | 6.52 | 0.11 | -0.35 | -0.56 | -0.14 | -3.32 | 1231.7 | **0.0009** | **0.0033** |
| *BSS-12-Temperance* | Q1 (<25th) | 54 | 4.51 | 0.08 | 4.77 | 0.13 | 0.25 | 0.01 | 0.49 | 2.083 | 1707.9 | **0.0374** | **0.0673** |
| ***BSS-12-Temperance*** | Q2 (25–50th) | 41 | 5.42 | 0.02 | 5.53 | 0.08 | 0.11 | -0.05 | 0.27 | 1.331 | 401.6 | 0.184 | 0.2366 |
| ***BSS-12-Temperance*** | Q3 (50–75th) | 71 | 5.89 | 0.01 | 5.75 | 0.1 | -0.13 | -0.33 | 0.06 | -1.377 | 883.2 | 0.1688 | 0.2251 |
| ***BSS-12-Temperance*** | Q4 (≥75th) | 68 | 6.56 | 0.03 | 6.39 | 0.07 | -0.18 | -0.31 | -0.05 | -2.676 | 1645.2 | **0.0075** | **0.0209** |
| ***BSS-12-Intellectual*** | Q1 (<25th) | 57 | 5.14 | 0.06 | 5.41 | 0.11 | 0.27 | 0.04 | 0.5 | 2.318 | 1400.6 | **0.0206** | **0.0412** |
| ***BSS-12-Intellectual*** | Q2 (25–50th) | 23 | 5.75 | 0 | 5.89 | 0.09 | 0.15 | -0.03 | 0.32 | 1.614 | 3057.7 | 0.1065 | 0.1598 |
| ***BSS-12-Intellectual*** | Q3 (50–75th) | 79 | 6.11 | 0.01 | 5.89 | 0.1 | -0.23 | -0.42 | -0.04 | -2.329 | 1705.4 | **0.02** | **0.0412** |
| ***BSS-12-Intellectual*** | Q4 (≥75th) | 75 | 6.78 | 0.02 | 6.53 | 0.06 | -0.25 | -0.38 | -0.12 | -3.805 | 508.6 | **0.0002** | **0.0007** |

**Table S9: Cognitive outcomes – imputed between-arm ANCOVA at $T_2$ (week 8)**

| *Task* | *Measure* | *Arm 1 n* | *Arm 2 n* | *Adj. Mean ± SE (Arm 1)* | *Adj. Mean ± SE (Arm 2)* | *F* | *p (raw)* | *p (FDR-BH)* | *Partial η²* |
|---|---|---|---|---|---|---|---|---|---|
| **2-Back** | Accuracy | 100 | 98 | 0.97 ± 0.00 | 0.97 ± 0.00 | 0.22 | 0.6394 | 0.9024 | 0 |

| | | | | | | | | | |
|---|---|---|---|---|---|---|---|---|---|
| **2-Back** | *Average Correct Reaction Time* | *100* | *98* | *761.67 ± 14.53* | *768.56 ± 13.41* | *0.127* | *0.722* | *0.9024* | *0* |
| **2-Back** | Average Incorrect Reaction Time | 33 | 35 | 775.21 ± 73.67 | 893.60 ± 66.62 | 1.701 | 0.1922 | 0.9024 | 0 |
| **2-Back** | Correct Time Over Incorrect Time | 33 | 35 | 0.97 ± 0.05 | 0.88 ± 0.05 | 1.721 | 0.1897 | 0.9024 | 0.001 |
| **Stroop** | Accuracy | 100 | 98 | 0.99 ± 0.01 | 0.98 ± 0.01 | 0.662 | 0.4158 | 0.9024 | 0 |
| **Stroop** | Congruent Accuracy | 99 | 98 | 0.99 ± 0.01 | 1.00 ± 0.01 | 0.214 | 0.6437 | 0.9024 | 0 |
| **Stroop** | Incongruent Accuracy | 96 | 98 | 0.99 ± 0.01 | 0.97 ± 0.01 | 0.758 | 0.3841 | 0.9024 | 0 |
| **Stroop** | Average Correct Reaction Time | 100 | 98 | 1234.98 ± 58.33 | 1216.81 ± 57.66 | 0.051 | 0.8217 | 0.913 | 0 |
| **Stroop** | Average Incongruent Correct Reaction Time | 96 | 98 | 1202.13 ± 59.01 | 1254.59 ± 54.85 | 0.443 | 0.5057 | 0.9024 | 0 |
| **Stroop** | Average Congruent Correct Reaction Time | 99 | 98 | 1139.93 ± 55.67 | 1140.34 ± 55.23 | 0 | 0.9958 | 0.9958 | 0 |

**Table S10: Cognitive outcomes – imputed replication analysis - arm 2 (within-subject) at (week 16 vs week 8) $T_3$ vs $T_2$ t-test**

| *Task* | Measure | n | $T_2$ Mean ± SEM | $T_3$ Mean ± SEM | Δ ($T_3$−$T_2$) | 95% CI Low | 95% CI High | t | df | p (raw) | p (FDR-BH) | Cohen $d_z$ |
|---|---|---|---|---|---|---|---|---|---|---|---|---|
| ***2-Back*** | Accuracy | 98 | 0.97 ± 0.01 | 0.97 ± 0.01 | 0 | -0.01 | 0.01 | -0.8 | 2387.399 | 0.4239 | 0.5299 | -0.08 |
| ***2-Back*** | Average Correct Reaction Time | 98 | 775.51 ± 16.37 | 716.90 ± 19.81 | -53.2 | -89.24 | -17.15 | -2.895 | 1715.113 | **0.0038** | **0.0096** | -0.29 |
| ***2-Back*** | Average Incorrect Reaction Time | 38 | 914.63 ± 76.33 | 808.50 ± 68.99 | -117.06 | -294.97 | 60.86 | -1.29 | 6554.271 | 0.1972 | 0.3286 | -0.21 |
| ***2-Back*** | Correct Time Over Incorrect Time | 38 | 0.86 ± 0.04 | 0.88 ± 0.05 | 0.02 | -0.1 | 0.15 | 0.376 | 5010.877 | 0.707 | 0.707 | 0.06 |
| ***Stroop*** | Accuracy | 98 | 0.98 ± 0.01 | 0.99 ± 0.00 | 0.01 | -0.01 | 0.04 | 1.302 | 132618.405 | 0.193 | 0.3286 | 0.13 |
| ***Stroop*** | Congruent Accuracy | 98 | 0.99 ± 0.00 | 1.00 ± 0.00 | 0 | -0.01 | 0.01 | 0.688 | 28127.26 | 0.4914 | 0.546 | 0.07 |
| ***Stroop*** | Incongruent Accuracy | 98 | 0.97 ± 0.01 | 0.99 ± 0.00 | 0.02 | -0.01 | 0.05 | 1.169 | 85581.45 | 0.2424 | 0.3463 | 0.12 |
| ***Stroop*** | Average Correct Reaction Time | 98 | 1278.04 ± 63.82 | 1026.12 ± 46.88 | -240.09 | -337.13 | -143.05 | -4.85 | 7132.206 | **0** | **0** | -0.49 |
| ***Stroop*** | Average Incongruent Correct Reaction Time | 98 | 1297.18 ± 65.16 | 1065.25 ± 54.81 | -211.08 | -322.23 | -99.92 | -3.722 | 12473.621 | **0.0002** | **0.0007** | -0.38 |
| ***Stroop*** | Average Congruent Correct Reaction Time | 98 | 1208.15 ± 64.39 | 963.90 ± 43.34 | -228.52 | -341.66 | -115.38 | -3.959 | 6410.696 | **0.0001** | **0.0004** | -0.4 |

**Table S11: Cognitive outcomes – imputed sustainability analysis - arm 1 (within-subject) at $T_3$ vs $T_2$ (week 16 vs week 8) t-test**

| *Task* | *Measure* | *n* | *$T_2$ Mean ± SEM* | *$T_3$ Mean ± SEM* | *Δ ($T_3$-$T_2$)* | *95% CI Low* | *95% CI High* | *t* | *df* | *p (raw)* | *p (FDR-BH)* | *Cohen $d_z$* |
|---|---|---|---|---|---|---|---|---|---|---|---|---|
| **2-Back** | Accuracy | 100 | 0.97 ± 0.00 | 0.95 ± 0.01 | -0.02 | -0.03 | -0.01 | -3.038 | 382.5429 | **0.0025** | **0.0051** | -0.3 |
| **2-Back** | Average Correct Reaction Time | 100 | 748.60 ± 16.88 | 690.06 ± 13.46 | -58.56 | -89.51 | -27.6 | -3.719 | 424.5215 | **0.0002** | **0.0011** | -0.37 |

| | | | | | | | | | | | | |
|---|---|---|---|---|---|---|---|---|---|---|---|---|
| **2-Back** | Average Incorrect Reaction Time | 48 | 766.69 ± 61.84 | 818.75 ± 49.74 | 37.63 | -118.37 | 193.64 | 0.473 | 1570.0314 | 0.6362 | 0.6362 | 0.07 |
| **2-Back** | Correct Time Over Incorrect Time | 48 | 0.95 ± 0.05 | 0.88 ± 0.04 | -0.07 | -0.2 | 0.06 | -1.003 | 988.8867 | 0.316 | 0.395 | -0.14 |
| **Stroop** | Accuracy | 100 | 0.99 ± 0.01 | 0.97 ± 0.01 | -0.02 | -0.05 | 0.01 | -1.243 | 557.1609 | 0.2145 | 0.344 | -0.12 |
| **Stroop** | Congruent Accuracy | 99 | 0.99 ± 0.01 | 0.98 ± 0.01 | -0.01 | -0.05 | 0.02 | -0.791 | 480.8727 | 0.4295 | 0.4773 | -0.08 |
| **Stroop** | Incongruent Accuracy | 97 | 0.98 ± 0.01 | 0.96 ± 0.02 | -0.02 | -0.06 | 0.01 | -1.175 | 444.6355 | 0.2408 | 0.344 | -0.12 |
| **Stroop** | Average Correct Reaction Time | 100 | 1171.63 ± 72.39 | 1009.66 ± 51.02 | -164.64 | -252.98 | -76.3 | -3.657 | 1062.3039 | **0.0003** | **0.0011** | -0.37 |
| **Stroop** | Average Incongruent Correct Reaction Time | 97 | 1150.87 ± 63.87 | 1012.65 ± 56.00 | -187.31 | -298.43 | -76.18 | -3.31 | 663.5653 | **0.001** | **0.0025** | -0.34 |
| **Stroop** | Average Congruent Correct Reaction Time | 99 | 1082.04 ± 66.98 | 924.26 ± 40.68 | -160.21 | -247.44 | -72.98 | -3.602 | 2387.9823 | **0.0003** | **0.0011** | -0.36 |

**Table S12: Cognitive outcomes – imputed clinical-impact analysis – arm 1 and arm 2 pooled over intervention period and stratified by clinical severity - post vs pre t-test**

| *Task* | *Measure* | *Quartile* | *n* | *Pre Mean* | *Pre SEM* | *Post Mean* | *Post SEM* | *Δ (Post-Pre)* | *95% CI Low* | *95% CI High* | *t* | *df* | *p (raw)* | *p (FDR-BH)* |
|---|---|---|---|---|---|---|---|---|---|---|---|---|---|---|
| ***2-Back*** | *Accuracy* | *Q1 (<25%)* | *49* | *0.86* | *0.01* | *0.91* | *0.01* | *0.05* | *0.02* | *0.08* | *3.216* | *18406.8* | ***0.0013*** | ***0.0058*** |
| ***2-Back*** | Accuracy | Q2 (25–50%) | 49 | 0.99 | 0 | 0.98 | 0 | -0.01 | -0.02 | 0 | -1.831 | 1020 | 0.0673 | 0.1282 |
| ***2-Back*** | Accuracy | Q3 (50–75%) | 49 | 1 | 0 | 1 | 0 | 0 | -0.01 | 0 | -1.386 | 612.6 | 0.1662 | 0.2891 |
| ***2-Back*** | Accuracy | Q4 (≥75%) | 49 | 1 | 0 | 0.99 | 0 | -0.01 | -0.02 | 0 | -1.672 | 296.6 | 0.0956 | 0.1738 |
| ***2-Back*** | Average Correct Reaction Time | Q1 (<25%) | 49 | 602.8 | 8.92 | 617.68 | 16.05 | 14.89 | -18.32 | 48.09 | 0.879 | 3618.5 | 0.3795 | 0.506 |
| ***2-Back*** | Average Correct Reaction Time | Q2 (25–50%) | 49 | 733.48 | 3.96 | 702.26 | 16.99 | -31.22 | -63.95 | 1.51 | -1.872 | 792.2 | 0.0615 | 0.123 |
| ***2-Back*** | Average Correct Reaction Time | Q3 (50–75%) | 49 | 825.99 | 4.23 | 765.74 | 24.47 | -60.26 | -108.69 | -11.82 | -2.439 | 2952.1 | **0.0148** | **0.0426** |
| ***2-Back*** | Average Correct Reaction Time | Q4 (≥75%) | 49 | 999.34 | 18.02 | 842.42 | 29.22 | -156.92 | -221.64 | -92.2 | -4.755 | 1857.1 | **0** | **0** |
| ***2-Back*** | Average Incorrect Reaction Time | Q1 (<25%) | 17 | 567.89 | 22.34 | 630.01 | 44.67 | 62.12 | -37.06 | 161.3 | 1.23 | 613.4 | 0.2191 | 0.3652 |
| ***2-Back*** | Average Incorrect Reaction Time | Q2 (25–50%) | 17 | 714.11 | 7.28 | 747.93 | 77.9 | 33.82 | -115.51 | 183.16 | 0.444 | 15978.1 | 0.6571 | 0.7667 |
| ***2-Back*** | Average Incorrect Reaction Time | Q3 (50–75%) | 16 | 888.05 | 20.51 | 859.22 | 113.36 | -28.83 | -247.24 | 189.58 | -0.259 | 2564.9 | 0.7958 | 0.8377 |

| | | | | | | | | | | | | | | |
|---|---|---|---|---|---|---|---|---|---|---|---|---|---|---|
| ***2-Back*** | Average Incorrect Reaction Time | Q4 (≥75%) | 16 | 1535.81 | 100.85 | 894.78 | 115.76 | -641.04 | -876.35 | -405.72 | -5.344 | 1467 | **0** | **0** |
| ***2-Back*** | Correct Time Over Incorrect Time | Q1 (<25%) | 17 | 0.54 | 0.03 | 0.87 | 0.07 | 0.34 | 0.19 | 0.48 | 4.493 | 486.7 | **0** | **0.0001** |
| ***2-Back*** | Correct Time Over Incorrect Time | Q2 (25–50%) | 17 | 0.79 | 0.01 | 0.87 | 0.08 | 0.08 | -0.07 | 0.22 | 0.997 | 881.8 | 0.3192 | 0.4403 |
| ***2-Back*** | Correct Time Over Incorrect Time | Q3 (50–75%) | 16 | 0.92 | 0.01 | 0.94 | 0.05 | 0.01 | -0.09 | 0.12 | 0.231 | 2173.9 | 0.8172 | 0.8381 |
| ***2-Back*** | Correct Time Over Incorrect Time | Q4 (≥75%) | 16 | 1.22 | 0.03 | 1.01 | 0.06 | -0.21 | -0.35 | -0.06 | -2.8 | 12118.2 | **0.0051** | **0.0186** |
| ***Stroop*** | Accuracy | Q1 (<25%) | 49 | 0.88 | 0.04 | 0.98 | 0.01 | 0.1 | 0.02 | 0.17 | 2.41 | 25483.5 | **0.016** | **0.0426** |
| ***Stroop*** | Accuracy | Q2 (25–50%) | 49 | 1 | 0 | 1 | 0 | 0 | -0.01 | 0.01 | -0.426 | 171.9 | 0.6708 | 0.7667 |
| ***Stroop*** | Accuracy | Q3 (50–75%) | 49 | 1 | 0 | 0.99 | 0 | -0.01 | -0.01 | 0 | -2.416 | 945.2 | **0.0159** | **0.0426** |
| ***Stroop*** | Accuracy | Q4 (≥75%) | 49 | 1 | 0 | 1 | 0 | 0 | -0.01 | 0 | -1.014 | 146.3 | 0.3122 | 0.4403 |
| ***Stroop*** | Congruent Accuracy | Q1 (<25%) | 49 | 0.96 | 0.02 | 0.98 | 0.01 | 0.02 | -0.02 | 0.06 | 1.061 | 2406059.1 | 0.2887 | 0.4403 |
| ***Stroop*** | Congruent Accuracy | Q2 (25–50%) | 49 | 1 | 0 | 1 | 0 | 0 | 0 | 0 | -0.1 | 192.2 | 0.9208 | 0.9208 |
| ***Stroop*** | Congruent Accuracy | Q3 (50–75%) | 49 | 1 | 0 | 1 | 0 | 0 | -0.01 | 0 | -1 | 1000000 | 0.3173 | 0.4403 |
| ***Stroop*** | Congruent Accuracy | Q4 (≥75%) | 48 | 1 | 0 | 1 | 0 | 0 | -0.01 | 0 | -0.336 | 171.8 | 0.7376 | 0.7974 |
| ***Stroop*** | Incongruent Accuracy | Q1 (<25%) | 48 | 0.85 | 0.05 | 0.97 | 0.02 | 0.12 | 0.02 | 0.22 | 2.441 | 11288 | **0.0147** | **0.0426** |
| ***Stroop*** | Incongruent Accuracy | Q2 (25–50%) | 48 | 1 | 0 | 1 | 0.01 | 0 | -0.02 | 0.01 | -0.432 | 154.8 | 0.6663 | 0.7667 |
| ***Stroop*** | Incongruent Accuracy | Q3 (50–75%) | 48 | 1 | 0 | 0.99 | 0.01 | -0.01 | -0.02 | 0 | -2.107 | 676.3 | **0.0355** | 0.0887 |
| ***Stroop*** | Incongruent Accuracy | Q4 (≥75%) | 47 | 1 | 0 | 0.99 | 0 | -0.01 | -0.01 | 0 | -1.144 | 120.5 | 0.2548 | 0.4076 |
| ***Stroop*** | Average Correct Reaction Time | Q1 (<25%) | 49 | 781.22 | 13.15 | 845.93 | 36.06 | 64.7 | -1.24 | 130.64 | 1.924 | 5407.6 | 0.0544 | 0.121 |
| ***Stroop*** | Average Correct Reaction Time | Q2 (25–50%) | 49 | 978.35 | 8.01 | 1037.37 | 74.49 | 59.02 | -87.55 | 205.6 | 0.79 | 3224.8 | 0.4298 | 0.5546 |
| ***Stroop*** | Average Correct Reaction Time | Q3 (50–75%) | 49 | 1261.32 | 19.23 | 1037.81 | 60.26 | -223.51 | -341.75 | -105.27 | -3.712 | 607.6 | **0.0002** | **0.0011** |

| | | | | | | | | | | | | | | |
|---|---|---|---|---|---|---|---|---|---|---|---|---|---|---|
| ***Stroop*** | Average Correct Reaction Time | Q4 (≥75%) | 49 | 2125.49 | 83.93 | 1503.67 | 127.4 | -621.82 | -796.73 | -446.91 | -6.97 | 4199.4 | **0** | **0** |
| ***Stroop*** | Average Incongruent Correct Reaction Time | Q1 (<25%) | 48 | 770.84 | 12.69 | 836.69 | 34.27 | 65.85 | -0.58 | 132.28 | 1.944 | 2688.9 | 0.052 | 0.121 |
| ***Stroop*** | Average Incongruent Correct Reaction Time | Q2 (25–50%) | 48 | 988.93 | 9.59 | 1032.24 | 89.83 | 43.31 | -134.58 | 221.2 | 0.478 | 1566.8 | 0.633 | 0.7667 |
| ***Stroop*** | Average Incongruent Correct Reaction Time | Q3 (50–75%) | 48 | 1265.05 | 19.62 | 1132.35 | 69.23 | -132.7 | -271.43 | 6.03 | -1.876 | 1585.4 | 0.0608 | 0.123 |
| ***Stroop*** | Average Incongruent Correct Reaction Time | Q4 (≥75%) | 47 | 2222.91 | 89.85 | 1442.92 | 99.32 | -779.99 | -989.03 | -570.96 | -7.318 | 1921.1 | **0** | **0** |
| ***Stroop*** | Average Congruent Correct Reaction Time | Q1 (<25%) | 49 | 700.19 | 12.32 | 828.2 | 42.37 | 128.02 | 48.91 | 207.12 | 3.175 | 1339.2 | **0.0015** | **0.0061** |
| ***Stroop*** | Average Congruent Correct Reaction Time | Q2 (25–50%) | 49 | 929.38 | 9.66 | 913.17 | 44.03 | -16.21 | -105.26 | 72.84 | -0.358 | 590.9 | 0.7208 | 0.7974 |
| ***Stroop*** | Average Congruent Correct Reaction Time | Q3 (50–75%) | 49 | 1202.63 | 18.95 | 956.63 | 50.82 | -246 | -351.57 | -140.43 | -4.571 | 1394.3 | **0** | **0** |
| ***Stroop*** | Average Congruent Correct Reaction Time | Q4 (≥75%) | 48 | 2079.73 | 88.47 | 1418.41 | 128.55 | -661.32 | -857.86 | -464.79 | -6.596 | 6849.1 | **0** | **0** |

**Table S13: Physiological outcomes – imputed between-arm ANCOVA at $T_2$ (week 8)**

| *Measure* | *Arm 1 n* | *Arm 2 n* | *Adj. Mean ± SE (Arm 1)* | *Adj. Mean ± SE (Arm 2)* | *F* | *p (raw)* | *p (FDR-BH)* | *Partial η²* |
|---|---|---|---|---|---|---|---|---|
| ***Resting Heart Rate*** | 92 | 88 | 64.41 ± 0.49 | 65.25 ± 0.47 | 1.519 | 0.218 | 0.4359 | 0.001 |
| ***Deep RMSSD*** | 86 | 84 | 42.98 ± 1.88 | 42.55 ± 1.73 | 0.028 | 0.8664 | 0.8664 | 0 |

**Table S14: Physiological outcomes – imputed replication analysis - arm 2 (within-subject) at $T_3$ vs $T_2$ (week 16 vs week 8) t-test**

| *Measure* | **n** | **$T_2$ Mean ± SEM** | **$T_3$ Mean ± SEM** | **Δ ($T_3$−$T_2$)** | **95% CI Low** | **95% CI High** | **t** | **df** | **p (raw)** | **p (FDR-BH)** | **Cohen $d_z$** |
|---|---|---|---|---|---|---|---|---|---|---|---|
| ***Resting Heart Rate*** | 90 | 65.09 ± 0.87 | 64.48 ± 0.86 | -0.46 | -1.31 | 0.39 | -1.055 | 3402.0348 | 0.2917 | 0.5833 | -0.11 |
| ***Deep RMSSD*** | 87 | 42.02 ± 2.76 | 41.37 ± 2.99 | -0.23 | -3.38 | 2.92 | -0.143 | 576.5251 | 0.8864 | 0.8864 | -0.02 |

**Table S15: Physiological outcomes – imputed sustainability analysis - arm 1 (within-subject) at $T_3$ vs $T_2$ (week 16 vs week 8) t-test**

| *Measure* | *n (post)* | *$T_2$ Mean ± SEM* | *$T_3$ Mean ± SEM* | *Δ ($T_3$− $T_2$)* | *95% CI Low* | *95% CI High* | *t* | *df* | *p (raw)* | *p (FDR-BH)* | *Cohen $d_z$* |
|---|---|---|---|---|---|---|---|---|---|---|---|
| ***Resting Heart Rate*** | 95 | 64.34 ± 0.80 | 64.08 ± 0.81 | -0.17 | -1.14 | 0.79 | -0.355 | 628.5074 | 0.7224 | 0.7224 | -0.04 |
| ***Deep RMSSD*** | *89* | *42.56 ± 2.70* | *40.08 ± 2.33* | *-2.68* | *-5.98* | *0.63* | *-1.595* | *317.5586* | *0.1117* | *0.2234* | *-0.17* |

**Table S16: Physiological outcomes – imputed clinical-impact analysis – arm 1 and arm 2 pooled over intervention period and stratified by clinical severity - post vs pre t-test**

| *Measure* | *Quartile* | *n* | *Pre Mean* | *Pre SEM* | *Post Mean* | *Post SEM* | *Δ (Post-Pre)* | *95% CI Low* | *95% CI High* | *t* | *df* | *p (raw)* | *p (FDR-BH)* |
|---|---|---|---|---|---|---|---|---|---|---|---|---|---|
| ***Resting Heart Rate*** | Q1 (<25%) | 45 | 54.83 | 0.52 | 56.19 | 0.65 | 1.36 | 0.27 | 2.45 | 2.443 | 4962.9 | **0.0146** | 0.1169 |
| ***Resting Heart Rate*** | Q2 (25–50%) | 44 | 62.47 | 0.24 | 62.27 | 0.65 | -0.2 | -1.43 | 1.03 | -0.319 | 1255.7 | 0.7498 | 0.7498 |
| ***Resting Heart Rate*** | Q3 (50–75%) | 44 | 67.66 | 0.27 | 66.66 | 0.62 | -1 | -2.15 | 0.15 | -1.708 | 606.7 | 0.0881 | 0.1763 |
| ***Resting Heart Rate*** | Q4 (≥75%) | 45 | 74.5 | 0.49 | 73.06 | 0.89 | -1.43 | -2.97 | 0.1 | -1.837 | 2096.6 | 0.0663 | 0.1763 |
| ***Deep RMSSD*** | *Q1 (<25%)* | *42* | *19.69* | *0.77* | *21.5* | *1.14* | *1.81* | *-0.09* | *3.71* | *1.864* | *1431.5* | *0.0625* | *0.1763* |
| ***Deep RMSSD*** | Q2 (25–50%) | 42 | 31.08 | 0.51 | 33.25 | 2.49 | 2.18 | -2.54 | 6.89 | 0.905 | 3258.8 | 0.3656 | 0.4874 |
| ***Deep RMSSD*** | Q3 (50–75%) | 41 | 43.55 | 0.77 | 42.85 | 2.08 | -0.7 | -4.69 | 3.29 | -0.344 | 811.1 | 0.7313 | 0.7498 |
| ***Deep RMSSD*** | Q4 (≥75%) | 42 | 78.47 | 4.19 | 71.72 | 4.3 | -6.74 | -15.16 | 1.67 | -1.571 | 2166.7 | 0.1162 | 0.186 |

**Table S17: Consolidated summary of univariate analysis (non-imputed)**

| | *Outcomes* | *Between Arm (Week 8)* | *Replication* | *Follow-up* | *Greater effect in higher-severity participants* |
|---|---|---|---|---|---|
| ***Primary Psychological Outcomes*** | Anxiety (GAD-7) | ∘ | ** | ☑ Sustained | *** |
| | Mind Wandering (MWQ) | ** | *** | ☑ Sustained | *** |
| | Sleep (PSQI) | -- | -- | -- | *** |
| ***Primary Cognitive Outcomes*** | 2-Back Accuracy | -- | -- | -- | * |
| | 2-Back Reaction Time | -- | ∘ | -- | ** |
| | Stroop Accuracy | -- | -- | -- | ∘ |
| | Stroop Reaction Time | -- | ** | -- | *** |
| ***Primary Physiological Outcomes*** | Resting Heart Rate | -- | -- | -- | -- |
| | Heart Rate Variability | -- | -- | -- | -- |
| ***Secondary Psychological Outcomes*** | Social Connectedness (SCS) | ∘ | *** | ☑ Sustained | *** |
| | Rumination (RRQ) | ∘ | *** | ☑ Sustained | *** |
| | Stress (PSS) | -- | ∘ | -- | *** |
| | Quality of Life (QOL) | ∘ | ** | ☑ Sustained | *** |

∘ $p_{\text{uncorrected}} < 0.05$ * $p_{\text{FDR}}<0.05$ ** $p_{\text{FDR}} <0.01$ *** $p_{\text{FDR}} <0.001$

**Table S18: EFA factor scores - imputed**

| *Measure* | *Factor 1* | *Factor 2* |
|---|---|---|
| ***Perceived Stress (PSS)*** | **0.84** | -0.01 |
| ***Depression (PHQ-8)*** | **0.78** | 0.07 |
| ***Anxiety (GAD-7)*** | **0.74** | 0.06 |
| ***Rumination (RRQ)*** | **0.74** | 0.02 |
| ***Mind Wandering (MWQ)*** | **0.53** | -0.05 |
| ***Sleep Disturbance (PSQI)*** | 0.34 | 0.02 |
| ***Quality of Life (QOLS)*** | **-0.56** | 0.30 |
| ***Social Connectedness (SCS-R)*** | **-0.47** | **0.42** |
| ***Reflection (RRQ)*** | 0.15 | **0.42** |
| ***Temperance Strength (BSS-12)*** | -0.14 | **0.52** |
| ***Intellectual Strength (BSS-12)*** | 0.08 | **0.65** |
| ***Interpersonal Strength (BSS-12)*** | -0.01 | **0.75** |

**Table S19: Latent factor-based outcomes – imputed between-arm ANCOVA at $T_2$ (week 8)**

| *Measure* | *Arm 1 n* | *Arm 2 n* | *Adj. Mean ± SE (Arm 1)* | *Adj. Mean ± SE (Arm 2)* | *F* | *p (raw)* | *p (FDR-BH)* | *Partial η²* |
|---|---|---|---|---|---|---|---|---|
| ***Factor 1*** | 86 | 101 | -0.358 ± 0.056 | -0.102 ± 0.052 | 11.195 | **0.001** | **0.002** | 0.057 |
| ***Factor 2*** | 86 | 101 | -0.011 ± 0.057 | -0.079 ± 0.053 | 0.746 | 0.389 | 0.389 | 0.004 |

**Table S20: Latent factor-based outcomes – imputed replication analysis - arm 2 (within-subject) at $T_3$ vs $T_2$ (week 16 vs week 8) t-test**

| *Measure* | **n** | **$T_2$ Mean ± SEM** | **$T_3$ Mean ± SEM** | **Δ ($T_3$−$T_2$)** | **95% CI Low** | **95% CI High** | **t** | **df** | **p (raw)** | **p (FDR-BH)** | **Cohen $d_z$** |
|---|---|---|---|---|---|---|---|---|---|---|---|
| ***Factor 1*** | 101 | -0.089 ± 0.086 | -0.376 ± 0.092 | -0.288 | -0.399 | -0.176 | -5.106 | 100 | **0** | **0** | -0.508 |
| ***Factor 2*** | 101 | -0.128 ± 0.094 | -0.104 ± 0.106 | 0.024 | -0.135 | 0.183 | 0.297 | 100 | 0.7674 | 0.7674 | 0.03 |

**Table S21: Latent factor-based outcomes – imputed sustainability analysis - arm 1 (within-subject) at $T_3$ vs $T_2$ (week 16 vs week 8) t-test**

| *Measure* | *n* | *$T_2$ Mean ± SEM* | *$T_3$ Mean ± SEM* | *Δ ($T_3$− $T_2$)* | *95% CI Low* | *95% CI High* | *t* | *df* | *p (raw)* | *p (FDR-BH)* | *Cohen $d_z$* |
|---|---|---|---|---|---|---|---|---|---|---|---|
| ***Factor 1*** | 86 | 0.047 ± 0.102 | 0.144 ± 0.103 | 0.097 | -0.004 | 0.198 | 1.903 | 85 | 0.0604 | 0.1208 | 0.205 |
| ***Factor 2*** | 86 | -0.374 ± 0.104 | -0.398 ± 0.093 | -0.024 | -0.161 | 0.113 | -0.345 | 85 | 0.7311 | 0.7311 | -0.037 |

**Table S22: Latent factor-based outcomes – imputed clinical-impact analysis – arm 1 and arm 2 pooled over intervention period and stratified by clinical severity - post vs pre t-test**

| *Measure* | *Quartile* | | *Pre Mean* | *Pre SEM* | *Post Mean* | *Post SEM* | *Delta Mean* | *Delta SEM* | *95% CI Low* | *95% CI High* | *t* | *df* | *p (raw)* | *p (FDR-BH)* |
|---|---|---|---|---|---|---|---|---|---|---|---|---|---|---|
| ***Factor 1*** | Q1 (<25th) | 47 | -1.2359 | 0.0475 | -1.3620 | 0.0640 | -0.1261 | 0.0563 | -0.2394 | -0.0128 | -2.2405 | 46 | **0.0299** | 0.0599 |
| ***Factor 1*** | Q2 (25–50th) | 46 | -0.3995 | 0.0334 | -0.5007 | 0.0942 | -0.1012 | 0.0885 | -0.2795 | 0.0771 | -1.14328 | 45 | 0.2590 | 0.3453 |
| ***Factor 1*** | Q3 (50–75th) | 47 | 0.1949 | 0.0251 | -0.1841 | 0.0880 | -0.3789 | 0.0829 | -0.5457 | -0.2122 | -4.57352 | 46 | **0.0000** | **0.0001** |
| ***Factor 1*** | Q4 (≥75th) | 47 | 1.2040 | 0.0715 | 0.5425 | 0.1243 | -0.6615 | 0.1011 | -0.8651 | -0.4580 | -6.54339 | 46 | **0.0000** | **0.0000** |
| ***Factor 2*** | Q1 (<25th) | 47 | -1.2953 | 0.0766 | -1.0317 | 0.1143 | 0.2636 | 0.0938 | 0.0748 | 0.4523 | 2.81072 | 46 | **0.0072** | **0.0193** |
| ***Factor 2*** | Q2 (25–50th) | 46 | -0.2813 | 0.0246 | -0.2298 | 0.0691 | 0.0516 | 0.0649 | -0.0791 | 0.1823 | 0.7949 | 45 | 0.4308 | 0.4924 |
| ***Factor 2*** | Q3 (50–75th) | 47 | 0.3325 | 0.0307 | 0.3125 | 0.0914 | -0.0200 | 0.0929 | -0.2070 | 0.1669 | -0.21568 | 46 | 0.8302 | 0.8302 |
| ***Factor 2*** | Q4 (≥75th) | 47 | 1.0828 | 0.0426 | 0.8052 | 0.1479 | -0.2776 | 0.1440 | -0.5674 | 0.0122 | -1.92848 | 46 | 0.0600 | 0.0960 |

**Table S23: Partial correlation of EFA factor and psychological data deltas (imputed with outliers removed)**

| *Measure* | *Factor* | *n* | *r* | $R^2$ | *p (raw)* | *p (FDR-BH)* |
|---|---|---|---|---|---|---|
| ***GAD-7*** | Factor 1 | 158 | -0.2451 | 0.0601 | **0.0020** | **0.0123** |
| ***GAD-7*** | Factor 2 | 158 | 0.0491 | 0.0024 | 0.5430 | 0.6206 |
| ***PSQI*** | Factor 1 | 161 | -0.1004 | 0.0101 | 0.2080 | 0.3329 |
| ***PSQI*** | Factor 2 | 161 | -0.0951 | 0.0090 | 0.2330 | 0.3494 |
| ***MWQ*** | Factor 1 | 159 | -0.1555 | 0.0242 | 0.0519 | 0.1038 |
| ***MWQ*** | Factor 2 | 159 | 0.0768 | 0.0059 | 0.3389 | 0.4280 |
| ***SCS-R*** | Factor 1 | 161 | 0.3095 | 0.0958 | **0.0001** | **0.0017** |
| ***SCS-R*** | Factor 2 | 161 | -0.2496 | 0.0623 | **0.0015** | **0.0121** |
| ***RRQ-Rumination*** | Factor 1 | 160 | -0.1723 | 0.0297 | **0.0304** | 0.0912 |
| ***RRQ-Rumination*** | Factor 2 | 160 | 0.1568 | 0.0246 | **0.0491** | 0.1038 |
| ***PSS*** | Factor 1 | 161 | -0.1883 | 0.0354 | **0.0175** | 0.0699 |
| ***PSS*** | Factor 2 | 161 | -0.0100 | 0.0001 | 0.9005 | 0.9005 |
| ***QOLS*** | Factor 1 | 160 | 0.2914 | 0.0849 | **0.0002** | **0.0024** |
| ***QOLS*** | Factor 2 | 160 | -0.1385 | 0.0192 | 0.0826 | 0.1526 |

| | | | | | | |
|---|---|---|---|---|---|---|
| ***PHQ-8*** | Factor 1 | 157 | -0.1763 | 0.0311 | **0.0282** | 0.0912 |
| ***PHQ-8*** | Factor 2 | 157 | 0.0842 | 0.0071 | 0.2975 | 0.3966 |
| ***RRQ-Reflection*** | Factor 1 | 161 | 0.1932 | 0.0373 | **0.0147** | 0.0699 |
| ***RRQ-Reflection*** | Factor 2 | 161 | -0.1639 | 0.0268 | **0.0390** | 0.1038 |
| ***BSS-12-Interpersonal*** | Factor 1 | 161 | 0.0896 | 0.0080 | 0.2614 | 0.3690 |
| ***BSS-12-Interpersonal*** | Factor 2 | 161 | -0.1322 | 0.0175 | 0.0967 | 0.1658 |
| ***BSS-12-Temperance*** | Factor 1 | 160 | 0.0444 | 0.0020 | 0.5798 | 0.6325 |
| ***BSS-12-Temperance*** | Factor 2 | 160 | -0.1596 | 0.0255 | **0.0452** | 0.1038 |
| ***BSS-12-Intellectual*** | Factor 1 | 160 | -0.0531 | 0.0028 | 0.5077 | 0.6092 |
| ***BSS-12-Intellectual*** | Factor 2 | 160 | -0.0411 | 0.0017 | 0.6083 | 0.6348 |

**Table S24: Partial correlation of EFA factor and physiological data deltas (imputed with outliers removed)**

| ***Measure*** | ***Factor*** | ***n*** | ***r*** | ***$R^2$*** | ***p (raw)*** | ***p (FDR-BH)*** |
|---|---|---|---|---|---|---|
| ***Resting State HR*** | Factor 1 | 127 | 0.0790 | 0.0062 | 0.3810 | 0.3810 |
| ***Resting State HR*** | Factor 2 | 127 | -0.1716 | 0.0294 | 0.0557 | 0.2230 |
| ***Deep RMSSD*** | Factor 1 | 126 | -0.1207 | 0.0146 | 0.1819 | 0.3638 |
| ***Deep RMSSD*** | Factor 2 | 126 | 0.0929 | 0.0086 | 0.3050 | 0.3810 |

**Table S25: Partial correlation of EFA factor and cognition delta (imputed with outliers removed)**

| ***Measure*** | ***Factor*** | ***n*** | ***r*** | ***$R^2$*** | ***p (raw)*** | ***p (FDR-BH)*** |
|---|---|---|---|---|---|---|
| ***2-Back Accuracy*** | Factor 1 | 149 | -0.1108 | 0.0123 | 0.1814 | 0.7439 |
| ***Stroop Incongruent Accuracy*** | Factor 2 | 149 | -0.0328 | 0.0011 | 0.6936 | 0.7927 |
| ***2-Back Average Correct Reaction Time*** | Factor 1 | 149 | -0.0184 | 0.0003 | 0.8252 | 0.8252 |
| ***2-Back Average Correct Reaction Time*** | Factor 2 | 149 | 0.0586 | 0.0034 | 0.4807 | 0.7858 |
| ***Stroop Incongruent Accuracy*** | Factor 1 | 144 | -0.1116 | 0.0125 | 0.1860 | 0.7439 |
| ***Stroop Incongruent Accuracy*** | Factor 2 | 144 | 0.0743 | 0.0055 | 0.3797 | 0.7858 |
| ***2-Back Average Incongruent Correct Reaction Time*** | Factor 1 | 149 | 0.0427 | 0.0018 | 0.6072 | 0.7927 |
| ***2-Back Average Incongruent Correct Reaction Time*** | Factor 2 | 149 | 0.0572 | 0.0033 | 0.4911 | 0.7858 |

### Supplemental Figures

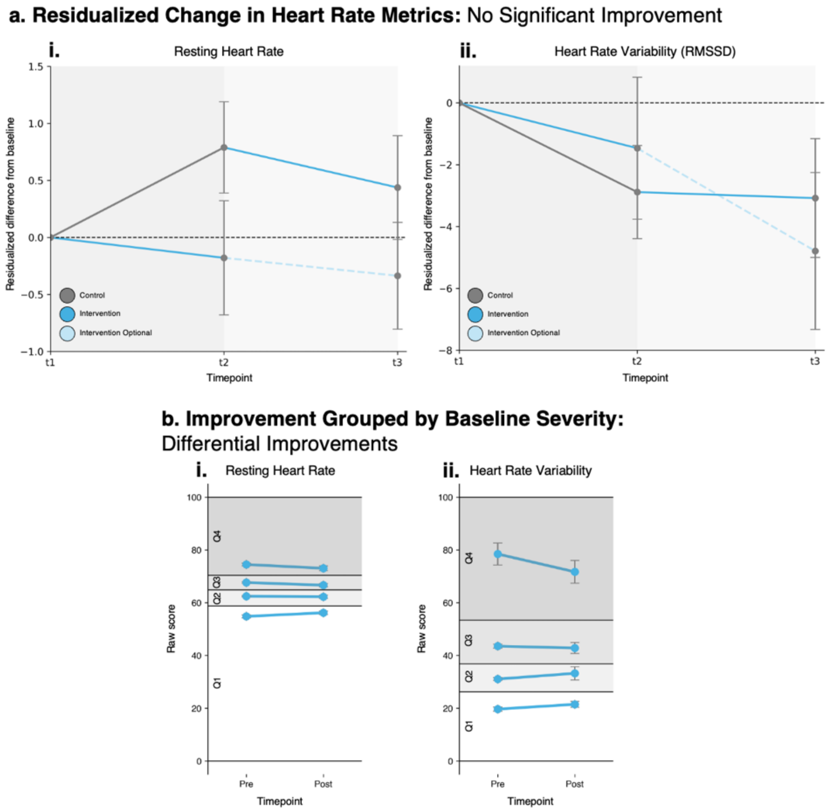


**Fig S1: Primary Physiological Outcomes.** Measures are reported in two different ways read left to right. Primarily, measures are reported as a time series and separated by arm. The y-axis represents residualized deviation-from-baseline score and the x-axis represents time point in the study (week 0, week 8, week 16). Lines are colored by study section: control period, intervention period, intervention optional period (sustainability analysis). Error bars represent standard error of the mean. [a] Resting heart rate decreases because of intervention while heart rate variability (HRV) decreases less in intervention than in control conditions. [b] There is no significant finding when stratified by quartiles.

**a. KMO**

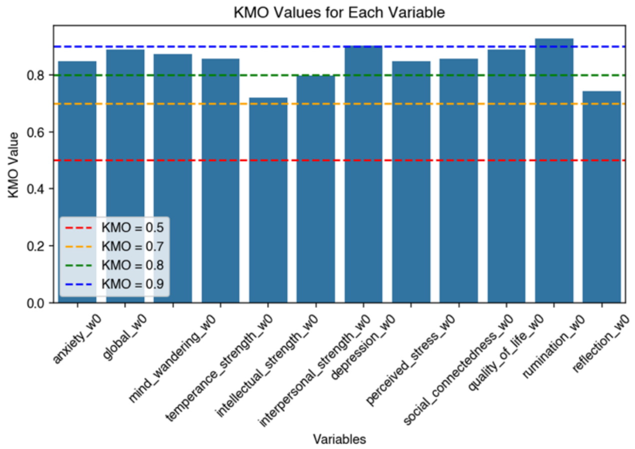


**Fig S2: Exploratory Factor Analysis.** [a] KMO for each variable show that EFA is allowable.
